# Establishing methods to monitor H5N1 influenza virus in dairy cattle milk

**DOI:** 10.1101/2024.12.04.24318491

**Authors:** Elyse Stachler, Andreas Gnirke, Kyle McMahon, Michael Gomez, Liam Stenson, Charelisse Guevara-Reyes, Hannah Knoll, Toni Hill, Sellers Hill, Katelyn S. Messer, Jon Arizti-Sanz, Fatinah Albeez, Elizabeth Curtis, Pedram Samani, Natalia Wewior, David H. O’Connor, William Vuyk, Sophia Khoury, Matthew K. Schnizlein, Nicole C. Rockey, Zachariah Broemmel, Michael Mina, Lawrence C. Madoff, Shirlee Wohl, Lorraine O’Connor, Catherine M. Brown, Al Ozonoff, Daniel J. Park, Bronwyn L. MacInnis, Pardis C. Sabeti

## Abstract

Highly Pathogenic Avian Influenza strain H5N1 has caused a multi-state outbreak among US dairy cattle, spreading across 15 states and infecting hundreds of herds since its onset. We rapidly developed and optimized PCR-based detection assays and sequencing protocols to support H5N1 molecular surveillance. Using 214 retail milk from 20 states for methods development, we found that H5N1 concentrations by digital PCR strongly correlated with qPCR cycle threshold (Ct) values, with dPCR exhibiting greater sensitivity. We also found that metagenomic sequencing after hybrid selection was best for higher concentration samples while amplicon sequencing performs best for lower concentrations. By establishing these methods, we were able to support the creation of a statewide surveillance program to test bulk milk samples monthly from all cattle dairy farms within Massachusetts, which remain negative to date. The methods, workflow, and recommendations described here provide a framework for others aiming to conduct H5N1 surveillance efforts.

## Introduction

Highly Pathogenic Avian Influenza strain H5N1 has caused large scale outbreaks in wild and domestic birds resulting in mass mortality, culling events, and economic losses (*1*). Viral spillover to mammals has become more frequent recently, including outbreaks with mammal-to-mammal transmission and sporadic human infections (*2*). In March 2024, H5N1 clade 2.3.4.4b was found in unpasteurized milk produced by infected dairy cattle in the US, the first confirmation of an outbreak that has grown to span more than 695 herds in 15 states as of October 24, 2024 (*3,4*). Phylogenetic analysis suggests the outbreak resulted from a single bird-to-cattle spillover event (*5*). The outbreak has subsequently spread through interstate transport of cattle, milking practices, and shared milking machinery and farm equipment (*6,7*). While confirmed human cases have thus far been sporadic and primarily with mild symptoms, the spread of H5N1 in cattle threatens the dairy industry and risks further adaptation to mammalian hosts, including humans.

This outbreak has highlighted the need for rapidly deployable H5N1 molecular surveillance capacity to detect infections, monitor viral spread and evolution, identify transmission routes, target interventions to protect agricultural assets and food supply, and prevent broader human transmission. Cow milk has emerged as an ideal sample source for H5N1 detection and surveillance during this outbreak; the virus is shed in high concentrations in milk, likely due to its affinity for infecting mammary gland epithelial cells (*8*). However, retail milk undergoes intense processing steps, including ultra-pasteurization and homogenization, with unknown effects on viral RNA quality.

While early reports detected the presence of H5N1 in pasteurized retail milk (*9-12*), optimized, robust, and scalable protocols are needed to routinely and reliably detect and characterize H5N1 for agricultural and public health surveillance. Using commercially available milk as a sample source, we evaluated and optimized methods for nucleic acid extraction, H5N1 RNA detection by digital and quantitative PCR, and library construction and sequencing approaches to produce near complete genomes. In partnership with the Massachusetts Departments of Agricultural Resources and Public Health, we then implemented quantitative detection protocols at scale to support routine statewide surveillance testing for H5N1 on all cattle milk-producing farms. Here we describe these methods and applications, along with considerations for implementation in other settings aiming to establish H5N1 detection and surveillance capacity.

## Results

### Validation, optimization, and performance of methods to detect H5N1 RNA in milk

To validate and optimize performance for molecular detection, we tested performance of an H5N1 assay using primers targeting the H5 subtype of the HA gene including sequences of the current outbreak strain (*13*) (H5_Taq) by both quantitative (qPCR) and digital PCR (dPCR). We optimized primer and probe concentrations using synthetic H5N1 RNA, selecting for optimal linearity, sensitivity, accuracy, precision, and qPCR assay efficiency (Figures A1 and A2).

Overall, the H5N1 assay displayed robust performance on both platforms, with dPCR outperforming qPCR in limit of detection (LOD) and precision. The LOD_90_ was 5 copies/μL by dPCR and 10 copies/μL by qPCR. Based on known concentrations of standard material, we found dPCR concentrations correlated well with qPCR Ct values (Figure 1D). In addition, dPCR exhibited lower coefficients of variations, ranging from 10.5-26.4% compared to 18.0-111.5% for qPCR evaluated for 2.5-25 copies/μL (Figure 1C). Both assays maintained linearity over their dynamic ranges (Figure 1A and 1B). Across all qPCR standard curves performed (n = 8), efficiencies ranged from 87-111% (Figure A3), with the 10^2^ copies standard concentration detected 93% of the time (Figure A5). This detection rate was lower than expected based on the LOD analysis, likely due to RNA standard degradation over time and highlighting the importance of standard material integrity for qPCR.

**Figure 1:**
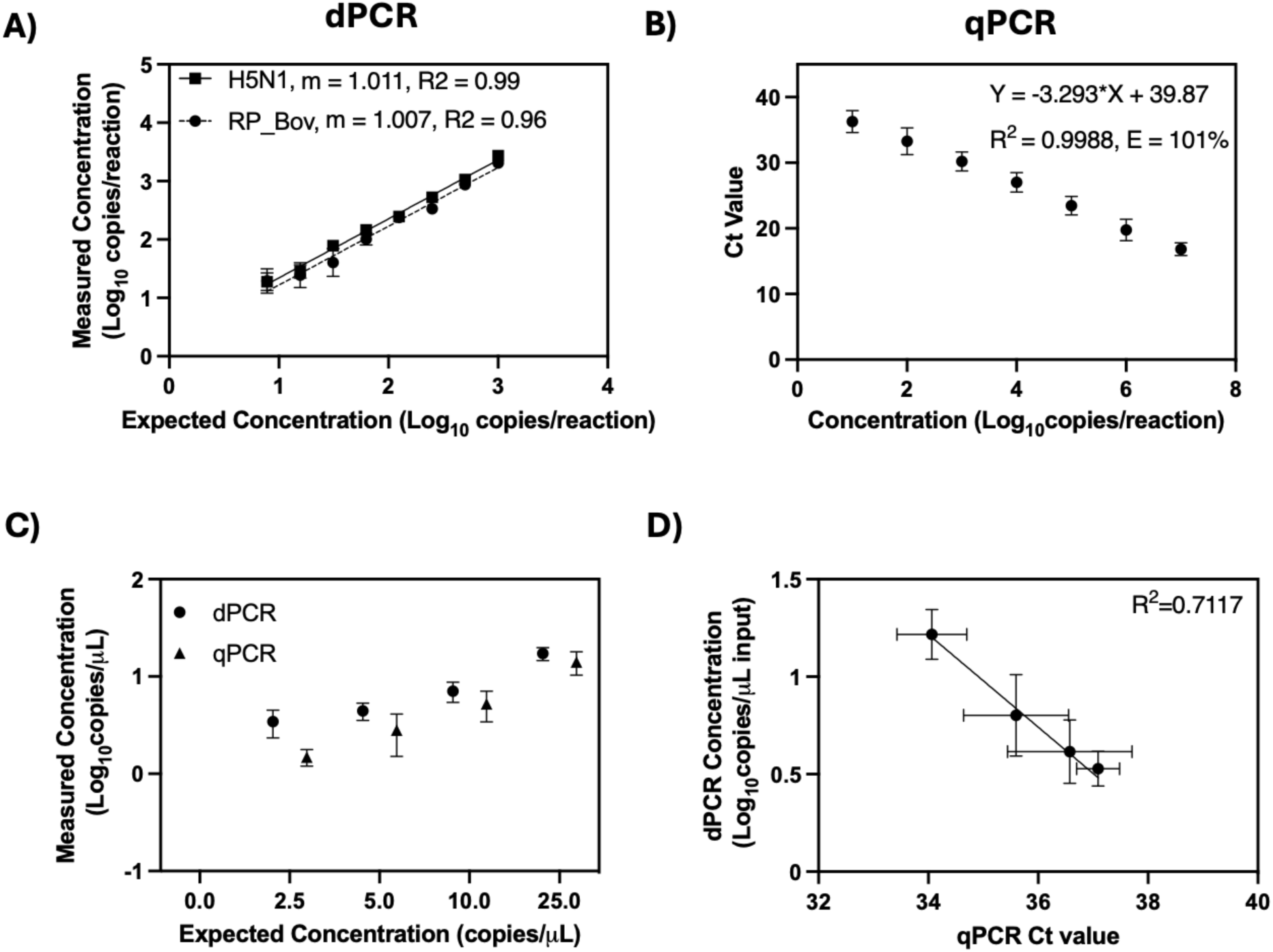
Validation and characterization of dPCR and qPCR H5N1 assays on synthetic spike-in samples. (A-B) Detection of (A) dPCR (H5_Taq and RP_Bov) and (B) qPCR (H5_Taq) assays using serial dilutions of synthetic H5N1 RNA standard material. For qPCR data, we combined and jointly analyzed all standard curve data from runs during retail milk testing. (C-D) Limit of Detection (LOD) analysis for (C) measured concentrations compared to expected concentrations for both qPCR and dPCR and (D) correlation of dPCR concentrations with qPCR Ct values. Fitted lines in (A) and (D) represent simple linear regression lines of best fit.

As a positive internal control for nucleic acid extraction in cattle milk, we designed a PCR assay targeting the bovine Ribonuclease P gene (both DNA and RNA; RP_Bov). By dPCR, linearity was maintained across all dilutions tested (Figure 1A) and the LOD_90_ was 10 copies/μL. Based on the superior performance of dPCR for the H5N1 target, the RP_Bov assay was not evaluated as a qPCR assay. Overall, all PCR assays performed well with minimal optimization.

We next evaluated pre-processing and extraction protocols to optimize sample preparation for subsequent H5N1 detection and sequencing. We tested two commercially available extraction kits, MagMAX Prime Viral/Pathogen (Prime) and MagMAX CORE (CORE), by spiking serial dilutions of synthetic H5N1 nucleic acid into milk. We tested milk with various fat contents and examined the effect of pre-centrifugation (at either 1200xg or 12,000xg) on outcomes. We also tested the MagMAX Wastewater kit (Wastewater) head-to-head with the CORE kit on a subset of eight retail milk samples previously found to be positive with CORE kit testing.

All three extraction kits demonstrated similar recovery of H5N1 from milk, with the CORE kit exhibiting slightly better performance. The CORE (Figure 2) and Prime (Figure A6) kits showed comparable results in terms of total recovery (down to ∼10^4^ H5N1 copies/mL milk) and linearity (R^2^_Prime,DNA_=0.93, R^2^_CORE,DNA_=0.96, and R^2^_CORE,RNA_=0.98). Milk fat content and pre-centrifugation exhibited no significant effect on target detection. As well, there was no significant difference in detection of H5N1 (p=0.20) or RP_Bov (p=0.17) using the Wastewater extraction kit on retail milk samples (Figure A7). We selected the CORE kit for ongoing testing given its low detection limit and slightly better detection of RP_Bov, as well as practical considerations including a manufacturer’s protocol for processing milk and kit availability.

**Figure 2:**
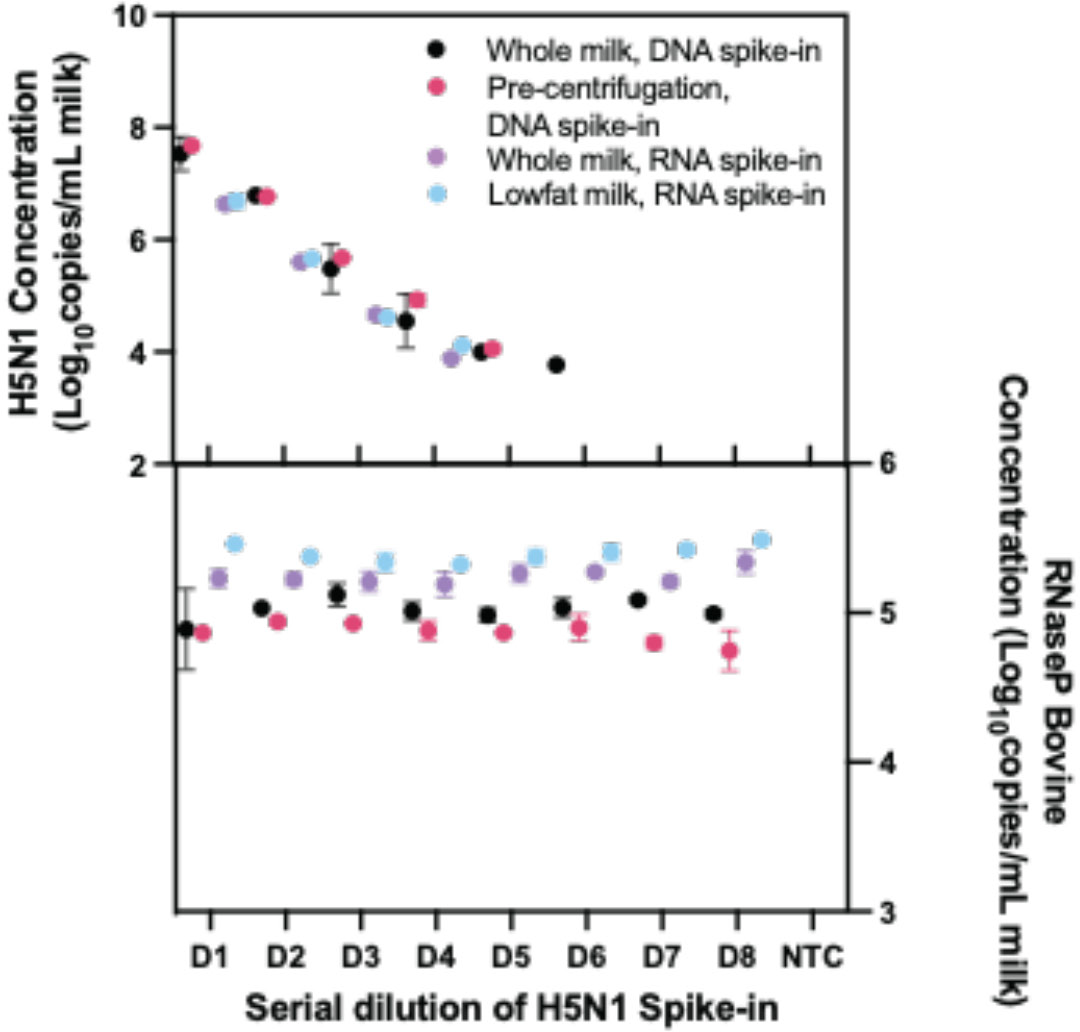
dPCR detection of H5N1 synthetic nucleic acid (top) and RNaseP Bovine (bottom) for the MagMAX CORE extraction kit. For direct extraction, we extracted 200μL of milk spiked with serial dilutions of H5N1 synthetic fragments. For pre-centrifugation, we centrifuged samples for 12000xg for 10 minutes following spike-in, after which we extracted 200μL.

### Detection of H5N1 in retail milk samples

To validate protocols on *in situ* H5N1 in milk, we sourced 214 retail milk cartons with diverse characteristics, including fat content and pasteurization processes, from 61 processing plants in 20 states (see Table, Figure 4A). Of these, 55 (26%) tested positive for presence of H5N1 RNA by dPCR, while 48 (22%) tested positive by qPCR. Positive samples were from processing plants in four states with reported H5N1 outbreaks (Colorado, Idaho, Michigan, and Texas). We also detected one positive sample by both dPCR and qPCR originating from a processing plant in Missouri, which has not reported H5N1 in cattle. Notably, the location of the processing plant reported on milk containers may or may not correspond to the state(s) in which the milk was initially collected, and this linkage is not publicly available.

**Figure 3:**
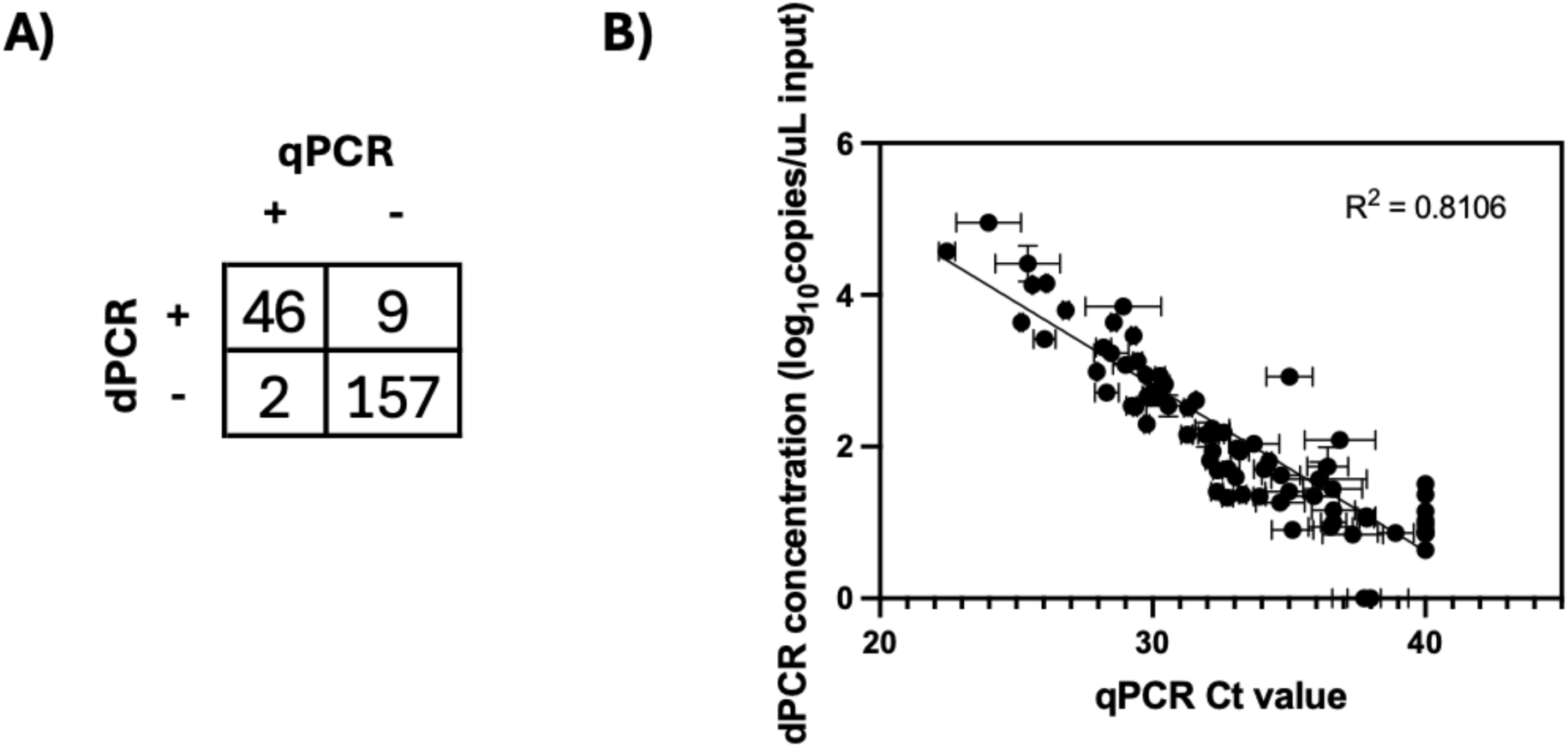
Comparison of dPCR and qPCR H5N1 testing on retail milk samples. (A) Agreement of positive and negative calls of milk samples between the two platforms. (B) Correlation of H5N1 measured by dPCR concentration compared with qPCR Ct value. For plotting purposes, samples not detected by dPCR were graphed with a dPCR concentration of 0 copies/μL while samples not detected by qPCR were graphed with a Ct value of 40.

**Figure 4:**
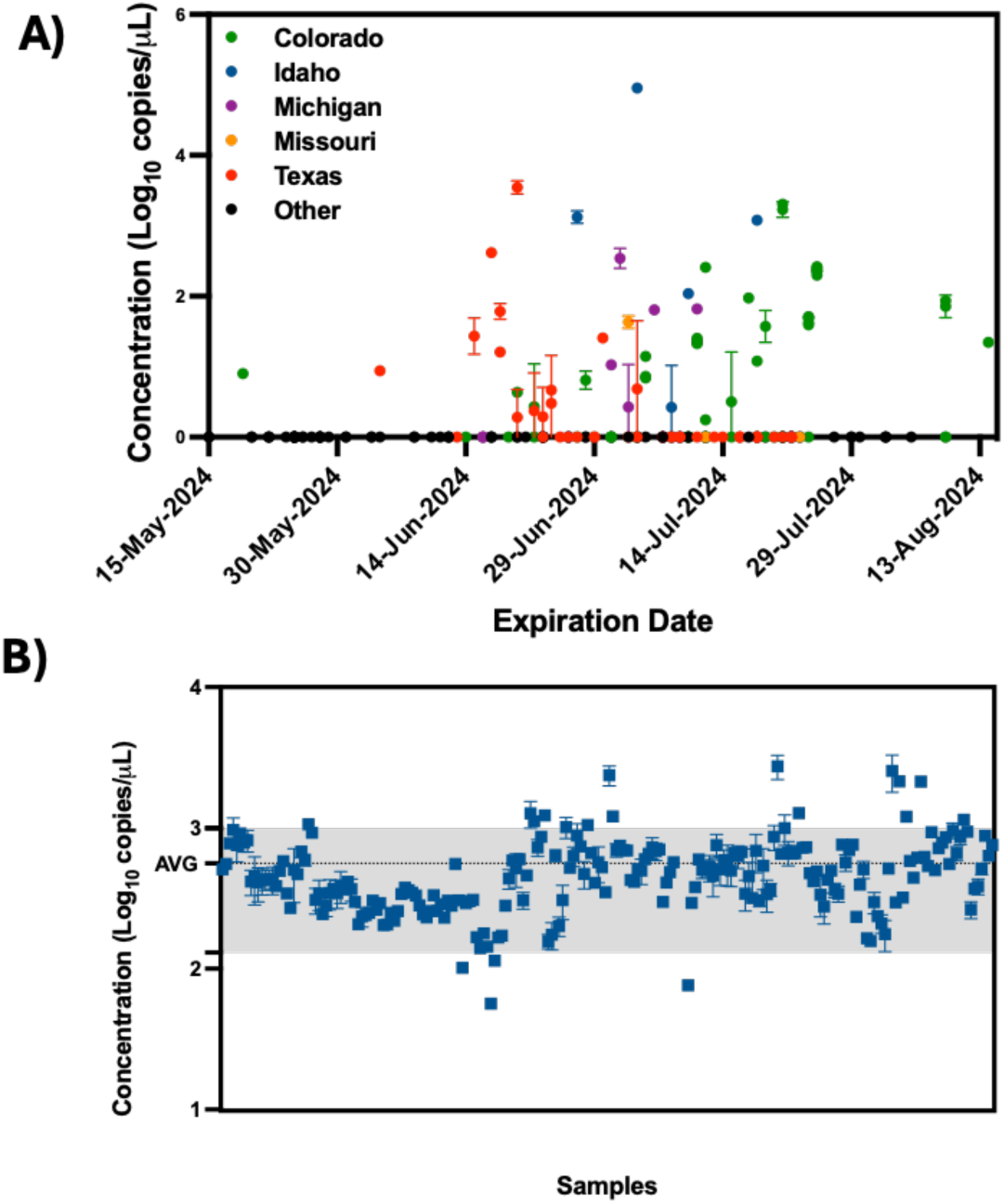
H5N1 and Bovine Ribonuclease P (RP_Bov) for all retail milk samples as measured by dPCR. A) The concentration of H5N1 as a function of processing state and expiration date. B) RP_Bov data for all samples. The gray-shaded region corresponds to the average RP_Bov concentration of all data plus and minus one standard deviation.

**Table:**
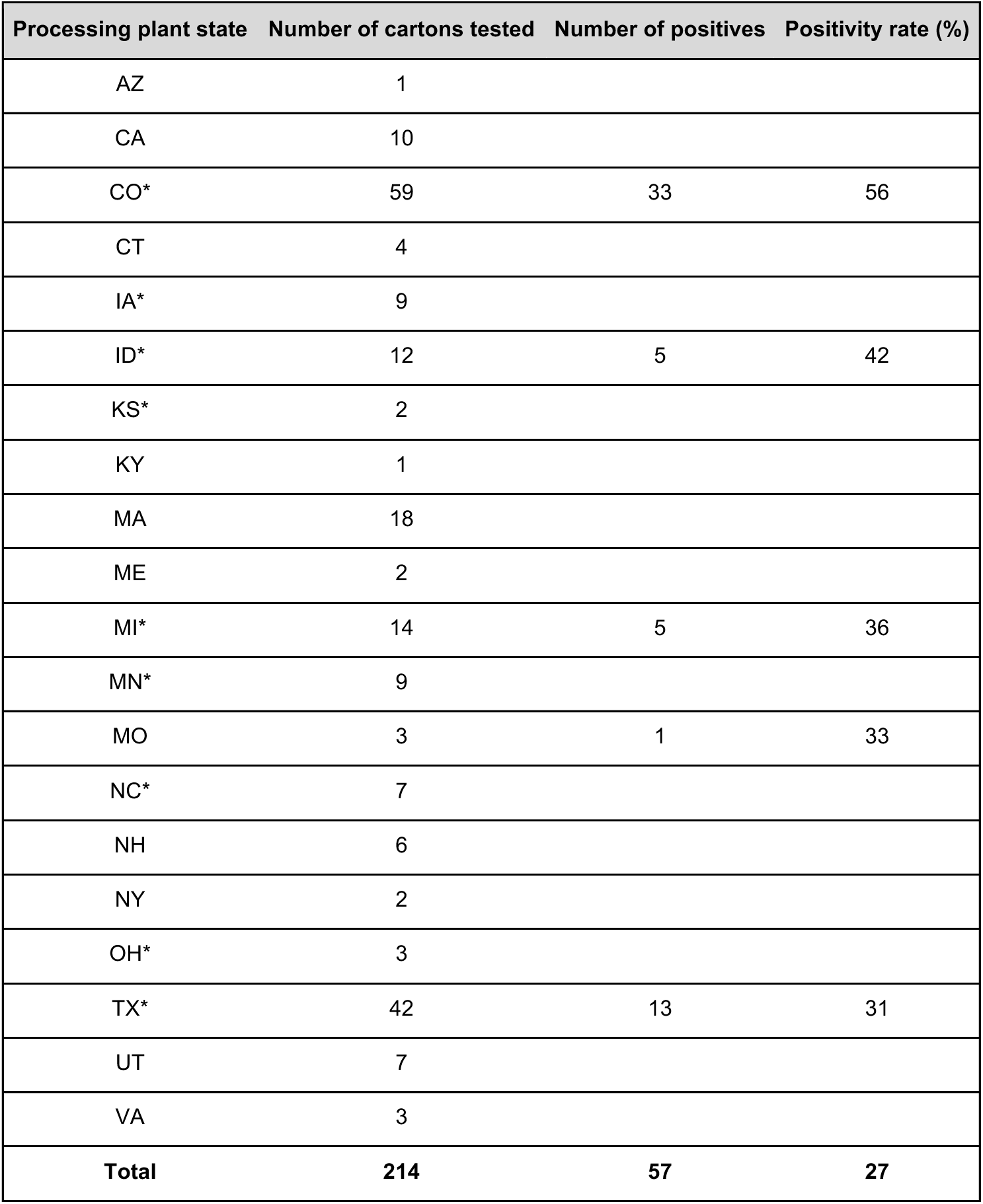
Breakdown of milk samples tested and their results by processing plant state. States designated with an asterisk (*) indicate states that had reported cases of H5N1 in cattle at the time of testing.

H5N1 detection and concentrations were strongly correlated between dPCR and qPCR platforms, with dPCR showing greater sensitivity. The platforms gave concordant positive/negative results for 95% (n=203/214) of samples (Figure 3A, see methods for thresholding details). Nine samples were positive only by dPCR, which may be due to the slightly enhanced LOD of the dPCR assay. Conversely, two samples were positive only by qPCR, possibly due to the more stringent thresholding criteria for dPCR. Further, H5N1 RNA dPCR concentrations correlated strongly with qPCR Ct values (R^2^=0.81, Figure 3B), suggesting the assay is robust on either platform.

We used the RP_Bov assay as an internal sample process control to confirm sample integrity and ensure proper collection and extraction, especially useful to interpret negative H5N1 results. RP_Bov concentrations averaged 560 copies/μL extract (Figure 4B), with 98% of samples falling within one standard deviation (we noted that four samples stored at 4°C for longer prior to processing than other samples fell below this lower limit). Thus, detection of RP_Bov below ∼100 copies/μL could be effectively used as a measure of milk sample and process integrity.

### Sequencing of H5N1 from retail milk

We attempted to recover genomes from 23 H5N1 positive retail milk samples, across a range of characteristics including virus concentration, milk type, and pasteurization process. To obtain higher H5N1 concentrations for library preparation, we first extracted, pooled, and concentrated ten samples from each milk container. Ultra-pasteurized samples exhibited significantly lower concentration factors compared to pasteurized samples as measured by H5N1 copy number (ultra-pasteurized_AVG_=4.6, pasteurized_AVG_=8.9, p=0.015, Figure A9). Despite being highly concentrated, samples showed no evidence of PCR inhibition by dPCR (p=0.89, Figure A10). The recovered RNA content and quality from these samples spanned a wide range as determined by H5N1 copies (14-9x10^4^ copies/μL), total RNA concentration (0.1-67.4 ng/μL), H5N1 copies/ng RNA (2-1.5x10^4^ copies/ng), and RIN (2-7.2) score (Table A7).

We evaluated three library construction methods to assess their efficacy in producing genomes across the range of H5N1 concentrations and pasteurization processes: untargeted metagenomic RNA sequencing (RNA-Seq), hybrid-selected RNA-Seq (hsRNA-Seq) enriched for human respiratory viruses including influenza A (albeit not explicitly H5N1) (*14*), and amplicon sequencing (Amp-Seq) of tiled 250-bp H5N1 PCR products (*15*). We produced near-complete (>70% assembly) H5N1 genomes from all 23 samples: 12 by hsRNA-Seq (≥80%) and 11 by Amp-Seq (≥74%). Hybrid selection greatly increased the chances of genome recovery for higher concentration extracts (>500 copies/μL), with hsRNA-Seq outperforming RNA-Seq for 11 of 12 samples. At lower concentrations, Amp-Seq resulted in the most complete genomes (Figure 5). Notably, we modified the PCR cycling conditions of a previously reported H5N1 Amp-Seq protocol (*15*), which resulted in improved amplicon generation and genome assemblies (Figure A11; see Methods). However, there was considerable variability in PCR efficiency across amplicons, with a small fraction of amplicons producing the vast majority of sequencing reads (Figure A12), thus still requiring a large amount of sequences per sample for complete genome assembly despite enrichment for specific PCR amplicons.

**Figure 5:**
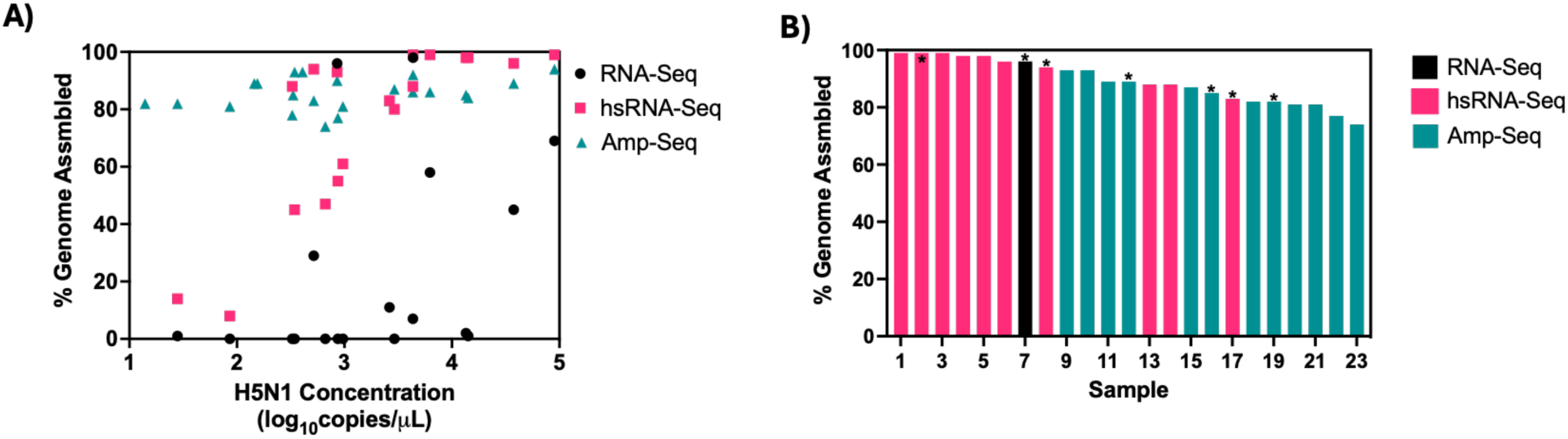
(A) Completeness of H5N1 genome assemblies generated by unbiased metagenomics (RNA-Seq), virus-enriched hybrid-selected metagenomics (hsRNA-Seq) and targeted H5N1 Amplicon Sequencing (Amp-Seq) as a function of H5N1 copies/μL RNA. (B): the most complete H5N1 assembly produced for each sample sorted by length and the underlying sequencing approach. Ultra-pasteurized samples are indicated by an (*) above the bar.

Phylogenetic analysis showed geographic clustering with other publicly available H5N1 genomes associated with the dairy cattle outbreak (Figure A13), suggesting the origin of the viruses was consistent with the US state of the processing plant of the milk. Notably, the positive originating from Missouri (which has no reports of H5N1 in cattle) clustered with samples from Texas and Michigan, likely pointing to the farm location the milk originated from despite being processed in a Missouri plant.

### Implementation at scale: statewide H5N1 surveillance of pre-pasteurized milk from Massachusetts farms

By establishing a robust workflow for detection and sequencing of H5N1 from milk, we were positioned to partner with the Massachusetts Departments of Agricultural Resources and of Public Health to support a mandatory surveillance program for H5N1 testing in milk from cattle dairy farms across the state. This program, launched in August 2024, was implemented preemptively in the absence of H5N1 detection in the state or surrounding region to confirm the absence of H5N1, and to serve as an early warning system should a local outbreak occur. State authorities worked with farms to collect samples from bulk milk tanks from all 95 cattle dairy farms across Massachusetts, initially within a 3 week period, followed by a rotating sampling schedule such that all farms were tested monthly. Based on our process validation using retail milk samples, we extracted bulk milk samples using the CORE extraction kit and performed dPCR for detection of H5_Taq and RP_Bov. While the surveillance program is currently ongoing, to date we have completed four rounds of statewide testing; H5N1 has not been detected in any sample from this surveillance program. The RP_Bov positive control has been routinely detected at similar levels to commercially available milk, providing confidence in the negative results obtained for H5N1 (Figure A14).

## Discussion

This study contributes validated methods for the whole workflow from sample to analyzed data for rapid deployment for potential future epidemiological studies and public health surveillance. We used retail milk products to optimize and validate methods for sample processing, detection, and sequencing of H5N1 in pasteurized milk, allowing us to implement these workflows at scale to support ongoing surveillance for H5N1 in pre-pasteurized milk from bulk tanks collected at the farm level. The workflow described here exhibits strong performance characteristics, while requiring minimal sample processing. We found direct nucleic acid extraction from milk regardless of fat content was efficient; with pre-centrifugation offering no increase in viral RNA recovery, in accordance with previous findings (*16,17*).

The H5N1 PCR assay performed well by both qPCR and dPCR and can be easily deployed on either platform. Both qPCR and dPCR have been widely adopted for viral surveillance, each with unique strengths: qPCR has a broader dynamic range of detection, higher throughput capabilities, lower cost per sample, and common instrument availability (*18,19*), while dPCR has been shown to have superior performance in clinical studies (*20, 21*), enhanced sensitivity (*18, 19, 21-23*), absolute quantification without reliance on a positive control standard (*18, 20*), robustness to PCR inhibitors (*19, 24*), and lower inter-laboratory variation (*25*). In cases where qPCR is used, dPCR can be useful for quantification of standards to ensure quality of materials (*20, 26*) to mitigate the effect of qPCR standard degradation and inaccuracy. The choice of PCR platform should weigh the features of each method, use case of the data, and the resources available to the laboratory.

Despite intense milk pre-processing (such as ultra-pasteurization), near-complete H5N1 genomes were readily recovered. Phylogenetic value of the genomes are limited in this context, as the original source farm(s) or state(s) are not provided on retail milk containers and the provided location of the processing plant may or may not correlate to the originating farm location. Using genome data, we verified that the positive sample obtained from a Missouri processing plant likely originated from a Texas farm. Notably, all other samples clustered geographically with publicly available H5N1 genomes originating from the state in which the milk was processed.

Our protocol development work enabled a partnership with state officials to perform preemptive mandatory H5N1 testing of raw milk from dairy farms across Massachusetts. To date the virus has not been detected in New England, likely due to being geographically far from where outbreaks have occurred thus far, the smaller size of the region’s dairy industry, and minimal interstate cattle transport compared to larger dairy industries (*27*). The prevalence of viral RNA in milk offers a unique surveillance mechanism to easily monitor lactating herds by testing pooled bulk milk tank samples, saving time and resources compared to individual cow testing. The sensitivity of our workflow allows for preemptive surveillance of H5N1 for the typical size of a Massachusetts cattle dairy farm (approximately 10,000 cows on 125 farms) (*28*). Based on our LOD of 10^4^ copies/mL milk, in order to detect 1 infected cow in a herd size of either 100 or 1000 cows, the infected cow would have to be shedding 10^6^ or 10^7^ H5N1 copies/mL milk, respectively. This is within the concentration range of live virus shed by infected cattle (10^4^-10^8.8^ TCID_50_/mL) (*8*).

Based on the methods testing and validation above, we offer guidance on establishing efficient, robust, and scalable H5N1 surveillance from bulk milk for implementation in molecular laboratory settings. For a detailed protocol and checklist, please refer to the guide provided in the Appendix. In brief, to obtain positive control material for protocol validation, look to source samples from outbreak-associated states by targeting milk cartons with USDA codes originating from processing plants in those states. The latest outbreak information (*4*) and USDA processing plant codes (*29*) can be easily accessed online. PCR assay performance should include linearity >90%, qPCR efficiency between 90-110%, and an LOD ≤ 10 copies/μL extract. Performance of the extraction method can be validated by spiking in synthetic nucleic acid into a negative milk sample, confirming that linearity > 90% and the process LOD ≤ 10^4^ copies/mL milk. For sequencing of H5N1 in milk, we recommend metagenomic sequencing after targeted hybrid selection for highly concentrated samples (>500 copies/μL) and sequencing of PCR amplicons for less concentrated or unconcentrated samples. Where permitted, near-complete genomes should be submitted to public databases. Finally, for unpasteurized milk testing, milk can be pasteurized in the laboratory per the USDA protocol (*30*) to limit risk of exposure to live virus while maintaining the molecular integrity of the sample for H5N1 detection and sequencing.

Enabling more labs to set up decentralized surveillance will allow us to stay ahead of current and future outbreaks of public concern. We hope the guidelines provided here can serve as a blueprint for rapid validation of new molecular detection methods and establishment of surveillance systems for the current H5N1 outbreak and beyond.

## Methods

### PCR assay design and characterization

For detection of H5N1 RNA, a previously published assay targeting the HA fragment of the H5N1 virus (denoted as H5_Taq) was used (*13*). Also, an internal extraction control assay was designed to target the bovine Ribonuclease P gene (RP_Bov). Digital PCR (dPCR) reactions were run on a Qiacuity One (5plex, Qiagen), while quantitative PCR (qPCR) assays were conducted on a QuantStudio 6 Flex (ThermoFisher). Additional details including primer sequences, kit information, primer and probe concentrations, and cycling conditions are reported in the Appendix. For dPCR, a sample had to have ≥ three positive partitions to be defined positive (*13, 31*). For qPCR, a sample was considered positive if 2 of 3 replicates amplified for at least one of the extraction replicates. The amplified Ct values were then averaged for subsequent analysis. PCR assay performance was evaluated using synthetic RNA of the H5N1 HA segment. The limit of detection (LOD_90_) was defined as the lowest concentration of target copies detected in at least 18/20 replicates. Linearity was evaluated from serial dilutions over the dynamic range for both PCR platforms. Finally, the standard curves of all qPCR runs were evaluated for overall assay efficiency.

### Extraction kit evaluation

Three commercially available extraction kits were evaluated for their potential to recover nucleic acid from a milk matrix. All kits chosen were bead-based, high-throughput kits compatible with the KingFisher Flex instrument (ThermoFisher). The MagMAX Prime kit and MagMAX CORE kit were tested by spiking serial dilutions of an 1800bp synthetic nucleic acid fragment of the H5N1 HA sequence. Parameters evaluated were fat content (whole vs. 2%), dilution with phosphate-buffered saline (PBS), and pre-centrifugation (either 12000xg for 10 minutes or 1200xg for 30 minutes). Finally, the MagMAX Wastewater kit was evaluated head-to-head with the MagMAX CORE kit on a subset of retail milk samples previously identified as positive through evaluation with the CORE kit. For this comparison, all samples were re-extracted with the CORE kit as well as processed with the Wastewater kit on the same day. All kits were used following the manufacturer’s instructions, using 200μL of milk as input.

### Sourcing and nucleic acid isolation of retail milk

Milk was purchased around greater Boston and obtained from states reported to be impacted by the H5N1 outbreak. To source milk from across the country, milk was purchased using mobile delivery apps by targeting local brands of milk and delivered to a collaborator. The collaborator aliquoted the milk into two falcon tubes, sealed in plastic bags, packed on ice, and shipped back to the Broad Institute overnight for processing. Upon receipt, one set of aliquots was stored at -80°C for preservation. The other aliquot proceeded to nucleic acid extraction.

All milk samples were extracted in duplicate using the MagMAX CORE kit on a KingFisher Flex following the manufacturer’s “Simple Workflow” for 200μL of milk input. A subset of positive samples were chosen to be evaluated by sequencing. To obtain enough RNA for sequencing, these samples were re-extracted by the CORE kit 10 times and subsequently concentrated using the RNA-Clean and Concentrator Kit (Zymo) using manufacturer protocols, including on column DNase treatment and adjusting the amount of binding buffer and ethanol to match the total elution volume and passing the entire volume in multiple loading steps through the column. Effects of inhibition of concentrated samples were evaluated by the dPCR H5_Taq assay spiking in two different volumes of template (1μL vs 2μL of input) and calculating total concentration per μL of extract.

### Surveillance of raw milk samples from Massachusetts farms

Bulk tank samples from all Massachusetts cattle dairy farms (n = 95) were collected by MDAR employees on a monthly basis starting on August 6, 2024 and delivered to the Broad Institute for processing. Samples were collected in 2 ounce plastic containers, typical for bulk tank sampling, and transported on ice to the lab for testing. Samples were immediately pasteurized onsite in a heated water bath by ensuring the internal temperature of the collection bottle reached 72°C for at least 15 seconds per USDA protocols (30). The samples were immediately placed on ice to cool and then proceeded through the same workflow as described above for nucleic acid extraction and subsequent dPCR analysis.

### Preparation of sequencing libraries

Three methods were evaluated for library construction: untargeted metagenomic RNA sequencing (RNA-Seq) using the xGen RNA library prep kit (IDT), hybrid-selected RNA-Seq (hsRNA-Seq) of xGen RNA-Seq libraries enriched using the Respiratory Virus Research Capture panel as bait (Twist Biosciences) with the Target Enrichment Standard Hybridization v2 kit following the manufacturer’s protocols, and amplicon sequencing (Amp-Seq) of tiled 250-bp H5N1 PCR products as described by Vuyk et al (*15*). For Amp-Seq, we used a modified thermoprofile from the original published protocol, namely shortening the annealing step and adding an extension step (Table A6). Full methods are described in detail in the Appendix. RNA samples for sequencing were run on an RNA 6000 Pico Bioanalyzer chip (Agilent) to determine total RNA concentration, size distribution, and RNA Integrity (RIN) scores. Water and a water extraction control served as blank negative controls for library construction. Pooled sequencing libraries were sequenced with paired-end 151-base reads on 300-cycle NextSeq 2000 cartridges (Illumina).

### Genomic analysis

Detailed methods for genomic analysis are available in the Appendix. Briefly, NextSeq sequencing runs were basecalled and demultiplexed using Picard using custom specified read structures to accommodate for the xGen library protocol, resulting in hard-trimmed reads containing only target sequences and obviating the need for post-alignment based trimming during consensus sequence generation. For each sequencing library, consensus influenza genomes were produced using a standard consensus generation pipeline utilized previously (*32-35*). The H5N1 Bovine/texas/24-029328-01/2024 reference genome (PP599462.1 through PP599469.1) was used as the reference for all assemblies. For each sample sequenced by multiple methods, we utilized the hsRNA-Seq genome if it recovered >75% of the genome, otherwise using the Amp-Seq genome. Phylogenetic analysis was performed by releasing successful genomes on NCBI Genbank, allowing automatic incorporation into the Moncla Lab/Nextstrain avian-flu builds for the cattle-associated outbreak. Sequence data is available at NCBI/INSDC under BioProject PRJNA1134696.

### Statistical analyses

All statistical analysis was completed in GraphPad Prism with statistical significance defined as p<0.05. Correlation between dPCR concentration and qPCR Ct value as well as tests of linearity were fit by simple linear regression. One-way ANOVA was used to compare effects of different conditions when multiple conditions were compared whereas a paired t-test was used when two conditions were being compared.

## Data Availability

All data produced in the present study are available upon reasonable request to the authors

## Acknowledgements

The authors thank the Boston Globe Media Partners, LLC staff for sourcing and donating local New England milk samples and John Rinn, Noreen Beckie, Kristen Koneschik, and Michael Butts for collecting US-based milk samples, and to other teammates/collaborators for helpful discussions.

## References

1. Peacock T, Moncla L, Dudas G, VanInsberghe D, Sukhova K, Lloyd-Smith JO, et al. The global H5N1 influenza panzootic in mammals. Nature. Forthcoming 2024. 10.1038/s41586-024-08054-z.

2. Graziosi G, Lupini C, Catelli E, Carnaccini S. Highly Pathogenic Avian Influenza (HPAI) H5 Clade 2.3.4.4b Virus Infection in Birds and Mammals. Animals (Basel). 2024 May 2;14(9):1372. doi: 10.3390/ani14091372. PMID: 38731377; PMCID: PMC11083745.

3. World Health Organization. Avian Influenza A(H5N1) – United States of America. 2024 April 9 [cited 2024 Oct 1]. https://www.who.int/emergencies/disease-outbreak-news/item/2024-DON512.

4. US Department of Agriculture Animal and Plant Health Inspection Service. HPAI Confirmed Cases in Livestock. 2024 Jul 3 [cited 2024 Dec 3]. https://www.aphis.usda.gov/livestock-poultry-disease/avian/avian-influenza/hpai-detections/hpai-confirmed-cases-livestock.

5. Nguyen T-Q, Hutter C, Markin A, Thomas M, Lantz K, Killian ML, et al. Emergence and interstate spread of highly pathogenic avian influenza A(H5N1) in dairy cattle. unpub. data, 10.1101/2024.05.01.591751.

6. Halwe NJ, Cool K, Breithaupt A, Schön J, Trujillo JD, Nooruzzaman M, et al. H5N1 clade 2.3.4.4b dynamics in experimentally infected calves and cows. Nature. Forthcoming 2024. 10.1038/s41586-024-08063-y.

7. Le Sage V, Campbell AJ, Reed DS, Duprex WP, Lakdawala SS. Persistence of Influenza H5N1 and H1N1 Viruses in Unpasteurized Milk on Milking Unit Surfaces. Emerg Infect Dis. 2024 Aug;30(8):1721–1723. doi: 10.3201/eid3008.240775. Epub 2024 Jun 24. PMID: 38914418; PMCID: PMC11286056.

8. Caserta LC, Frye EA, Butt SL, Laverack M, Nooruzzaman M, Covaleda LM, et al. Spillover of highly pathogenic avian influenza H5N1 virus to dairy cattle. Nature. 2024 Jul 25; 634, 669–676. 10.1038/s41586-024-07849-4.

9. Lauring AS, Martin ET, Eisenberg M, Fitzsimmons WJ, Salzman E, Leis AM, et al. Surveillance of H5 HPAI in Michigan using retail milk. unpub. data, 10.1101/2024.07.04.602115.

10. Spackman E, Jones DR, McCoig AM, Colonius TJ, Goraichuk IV, Suarez DL. Characterization of highly pathogenic avian influenza virus in retail dairy products in the US. J Virol. 2024 Jul 23;98(7):e0088124. doi: 10.1128/jvi.00881-24. Epub 2024 Jul 3. PMID: 38958444; PMCID: PMC11264905.

11. Suarez DL, Goraichuk PV, Killmaster L, Spackman E, Clausen NJ, Colonius TJ, et al. Testing of retail cheese, butter, ice cream and other dairy products for highly pathogenic avian influenza in the US. unpub. data, doi:10.1101/2024.08.11.24311811.

12. Tarbuck N, Jones J, Franks J, Kandeil A, DeBeauchamp J, Miller L, et al. Detection of A(H5N1) influenza virus nucleic acid in retail pasteurized milk. unpub. data, doi:10.21203/rs.3.rs-4572362/v1.

13. Wolfe MK, Duong D, Shelden B, Chan EMG, Chan-Herur V, Hilton S, et al. Detection of Hemagglutinin H5 Influenza A Virus Sequence in Municipal Wastewater Solids at Wastewater Treatment Plants with Increases in Influenza A in Spring, 2024. ES&T Letters. 2024 May 20;11(6), 526-532. doi: 10.1021/acs.estlett.4c00331.

14. Metsky HC, Siddle KJ, Gladden-Young A, Qu J, Yang DK, Brehio P, et al. Capturing sequence diversity in metagenomes with comprehensive and scalable probe design. Nat. Biotechnol. 2019 February 4; 37, 160–168. 10.1038/s41587-018-0006-x.

15. Vuyk WC, Lail A, Emmen I, Hassa N, Tiburcio PB, Newman C, et al. Whole genome sequencing of H5N1 from dairy products with tiled 250bp amplicons v1. 2024 Jun 5. protocols.io. 10.17504/protocols.io.kqdg322kpv25/v1.

16. Minsky BB, Kadam A, Tanner NA, Cantor E, Patton GC, New England Biolabs. Facilitating Purification and Detection of Viral Nucleic Acids from Milk. New England Biolabs. 2024 Jun. https://www.neb.com/en-us/-/media/nebus/files/application-notes/appnote_facilitating_purification_and_detection_of_viral_nucleicacids_from_milk.pdf?rev=d5777c15b56b470fb97e69ae6d855cfd&sc_lang=en-us.

17. Lail A, Vuyk WC, Emmen I, O’Connor D. RNA extraction from milk for HPAI surveillance v1. 2024 May 10. protocols.io. doi:10.17504/protocols.io.n2bvjn6obgk5/v1.

18. Tiwari A, Ahmed W, Oikarinen S, Sherchan SP, Heikinheimo A, Jiang G, et al. Application of digital PCR for public health-related water quality monitoring. Sci Total Environ. 2022 Sep 1;837:155663. doi: 10.1016/j.scitotenv.2022.155663. Epub 2022 May 4. PMID: 35523326.

19. Park S, Rana A, Sung W, Munir M. Competitiveness of Quantitative Polymerase Chain Reaction (qPCR) and Droplet Digital Polymerase Chain Reaction (ddPCR) Technologies, with a Particular Focus on Detection of Antibiotic Resistance Genes (ARGs). Applied Microbiology. 2021 Oct 2; 1(3):426–444. 10.3390/applmicrobiol1030028.

20. Kuypers J, Jerome KR. Applications of Digital PCR for Clinical Microbiology. J Clin Microbiol. 2017 May 23;55(6):1621–1628. doi: 10.1128/JCM.00211-17. Epub 2017 Mar 15. PMID: 28298452; PMCID: PMC5442518.

21. Liu C, Shi Q, Peng M, Lu R, Li H, et al. Evaluation of droplet digital PCR for quantification of SARS-CoV-2 Virus in discharged COVID-19 patients. Aging (Albany NY). 2020 Nov 1;12(21):20997–21003. doi: 10.18632/aging.104020. Epub 2020 Nov 1. PMID: 33136068; PMCID: PMC7695381.

22. Ahmed W, Simpson SL, Bertsch PM, Bibby K, Bivins A, Blackall LL, et al. Minimizing errors in RT-PCR detection and quantification of SARS-CoV-2 RNA for wastewater surveillance. Sci Total Environ. 2022 Jan 20;805:149877. doi: 10.1016/j.scitotenv.2021.149877. Epub 2021 Aug 25. PMID: 34818780; PMCID: PMC8386095.

23. Ahmed W, Smith WJM, Metcalfe S, Jackson G, Choi PM, Morrison M, et al. Comparison of RT-qPCR and RT-dPCR Platforms for the Trace Detection of SARS-CoV-2 RNA in Wastewater. ACS ES T Water. 2022 Nov 11;2(11):1871–1880. doi: 10.1021/acsestwater.1c00387. Epub 2022 Jan 28. PMID: 36380768; PMCID: PMC8848507.

24. Hinkle A, Greenwald HD, Metzger M, Thornton M, Kennedy LC, Loomis K, et al. Comparison of RT-qPCR and digital PCR methods for wastewater-based testing of SARS-CoV-2. unpub. data, doi:10.1101/2022.06.15.22276459.

25. Boxman ILA, Molin R, Persson S, Juréus A, Jansen CCC, Sosef NP, et al. An international inter-laboratory study to compare digital PCR with ISO standardized qPCR assays for the detection of norovirus GI and GII in oyster tissue. Food Microbiol. 2024 Jun;120:104478. doi: 10.1016/j.fm.2024.104478. Epub 2024 Jan 12. PMID: 38431324.

26. Park C, Lee J, Hassan ZU, Ku KB, Kim SJ, Kim HG, et al. Comparison of Digital PCR and Quantitative PCR with Various SARS-CoV-2 Primer-Probe Sets. J Microbiol Biotechnol. 2021 Mar 28;31(3):358–367. doi: 10.4014/jmb.2009.09006. PMID: 33397829; PMCID: PMC9705847.

27. Piore A. Traces of bird flu have made it into store-bought milk in New England, but at very low levels. The Boston Globe (MA). 2024 May 17. https://www.bostonglobe.com/2024/05/17/metro/inactivated-bird-flu-virus-found-in-milk-sold-in-massachusetts/.

28. USDA National Agricultural Statistics Service. 2023 Agricultural Statistics Annual Bulletin New England. 2024 Sep. https://www.nass.usda.gov/Statistics_by_State/New_England_includes/Publications/An nual_Statistical_Bulletin/2023/2023_NewEngland_Annual_Bulletin.pdf

29. Where Is My Milk From?. 2023 May 1 [cited 2024 Jul 31]. https://www.whereismymilkfrom.com.

30. Food and Drug Administration, Department of Health and Human Services. Code of Federal Regulations: Part 131–Milk and Cream. 1977 Mar 15. https://www.ecfr.gov/current/title-21/chapter-I/subchapter-B/part-131.

31. Huisman JS, Scire J, Caduff L, Fernandez-Cassi X, Ganesanandamoorthy P, Kull A, et al. Wastewater-Based Estimation of the Effective Reproductive Number of SARS-CoV-2. Environ Health Perspect. 2022 May 26;130(5):57011. doi: 10.1289/EHP10050. Epub 2022 May 26. PMID: 35617001; PMCID: PMC9135136.

32. Holmes EC, Dudas G, Rambaut A, Andersen KG. The evolution of Ebola virus: Insights from the 2013-2016 epidemic. Nature. 2016 Oct 13;538(7624):193-200. doi: 10.1038/nature19790. PMID: 27734858; PMCID: PMC5580494.

33. Lemieux JE, Siddle KJ, Shaw BM, Loreth C, Schaffner SF, Gladden-Young A, et al. Phylogenetic analysis of SARS-CoV-2 in Boston highlights the impact of superspreading events. Science. 2021 Feb 5;371(6529):eabe3261. doi: 10.1126/science.abe3261. Epub 2020 Dec 10. PMID: 33303686; PMCID: PMC7857412.

34. Wohl S, Metsky HC, Schaffner SF, Piantadosi A, Burns M, Lewnard JA, et al. Combining genomics and epidemiology to track mumps virus transmission in the United States. PLoS Biol. 2020 Feb 11;18(2):e3000611. doi: 10.1371/journal.pbio.3000611. PMID: 32045407; PMCID: PMC7012397.

35. Park D, Tomkins-Tinch C, Ye S, Jungreis I, Arenas FN, Shlyakhter I, et al. broadinstitute/viral-pipelines: v2.3.6.0. Zenodo. 2024 Nov. doi: 10.5281/zenodo.4306357

36. Metsky HC, Matranga CB, Wohl S, Schaffner SF, Freije CA, Winnicki SM, et al. Zika virus evolution and spread in the Americas. Nature. 2017 Jun 15;546(7658):411-415. doi: 10.1038/nature22402. Epub 2017 May 24. PMID: 28538734; PMCID: PMC5563848.

37. Bassano I, Ramachandran VK, Khalifa MS, Lilley CJ, Brown MR, van Aerle R, et al. Evaluation of variant calling algorithms for wastewater-based epidemiology using mixed populations of SARS-CoV-2 variants in synthetic and wastewater samples. Microb Genom. 2023 Apr;9(4):mgen000933. doi: 10.1099/mgen.0.000933. PMID: 37074153; PMCID: PMC10210938.

