## Supplemental Information for "Establishing methods to monitor H5N1 influenza virus in dairy cattle milk"

Author affiliations: Broad Institute of MIT and Harvard, Cambridge, Massachusetts, USA (E. Stachler, A. Gnirke, K. McMahon, M. Gomez, L. Stenson, C. Guevara-Reyes, H. Knoll, T. Hill, S. Hill, K. S. Messer, J. Arizti-Sanz, F. Albeez, E. Curtis, P. Samani, N. Wewior, A. Ozonoff, D. J. Park, B. L. MacInnis, P. C. Sabeti); University of Puerto Rico - Rio Piedras, San Juan, Puerto Rico, USA (C. Guevara-Reyes); Harvard University, Cambridge, Massachusetts, USA (P. Samani, B. MacInnis, P. C. Sabeti); University College London, London, United Kingdom (P. Samani); University of Wisconsin-Madison, Madison, Wisconsin, USA (D. O'Connor, W. Vuyk); The University of Texas at Austin, Austin, Texas, USA (S. Khoury); Michigan State University, East Lansing, MI, USA (M. K. Schnizlein); Duke University, Durham, NC, USA (N. C. Rockey, Z. Broemmel); Immune Observatory, Boston, Massachusetts, USA (M. Mina); Massachusetts

Department of Public Health, Boston, Massachusetts (L. C. Madoff, S. Wohl, C. M. Brown); University of Massachusetts Chan Medical School, Worcester, Massachusetts (L. C. Madoff); Brigham and Women's Hospital, Boston, Massachusetts, USA (S. Wohl); Massachusetts Department of Agricultural Resources, Boston, Massachusetts (L. O'Connor); Boston Children's Hospital, Boston, Massachusetts, USA (A. Ozonoff); Harvard Medical School, Boston, Massachusetts, USA (A. Ozonoff); Howard Hughes Medical Institute, Chevy Chase, Maryland, USA (P. C. Sabeti)

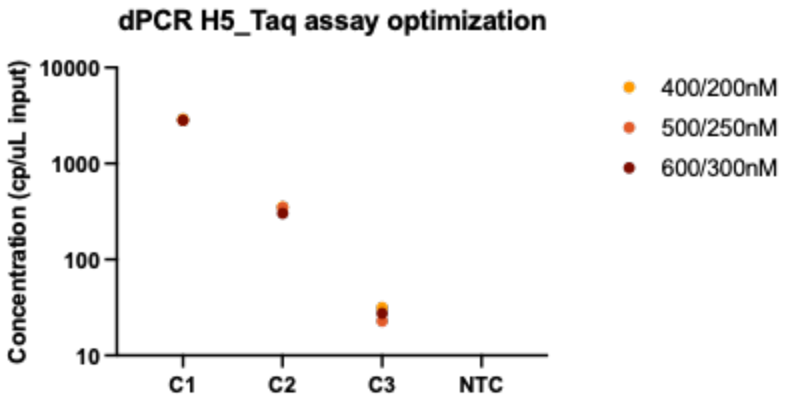


Figure A1: Optimization of primer and probe concentrations for H5_Taq dPCR assay using synthetic nucleic acid targets over three orders of magnitude (C1-C3). Based on this, 400/200nM final primer/probe concentration was selected.

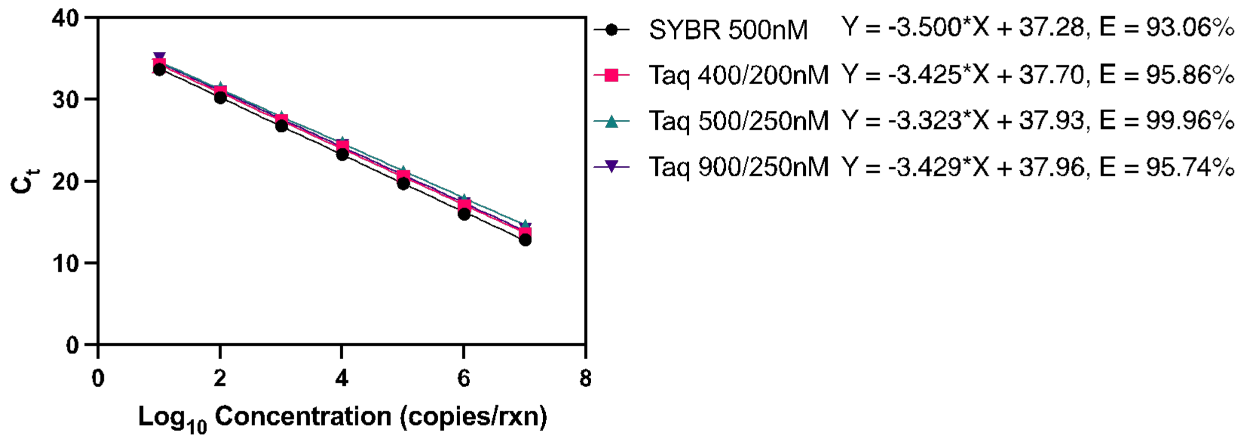


Figure A2: Optimization of primer and probe concentrations for H5_Taq qPCR assay. Based on this, 500/250nM final primer/probe concentration was selected.

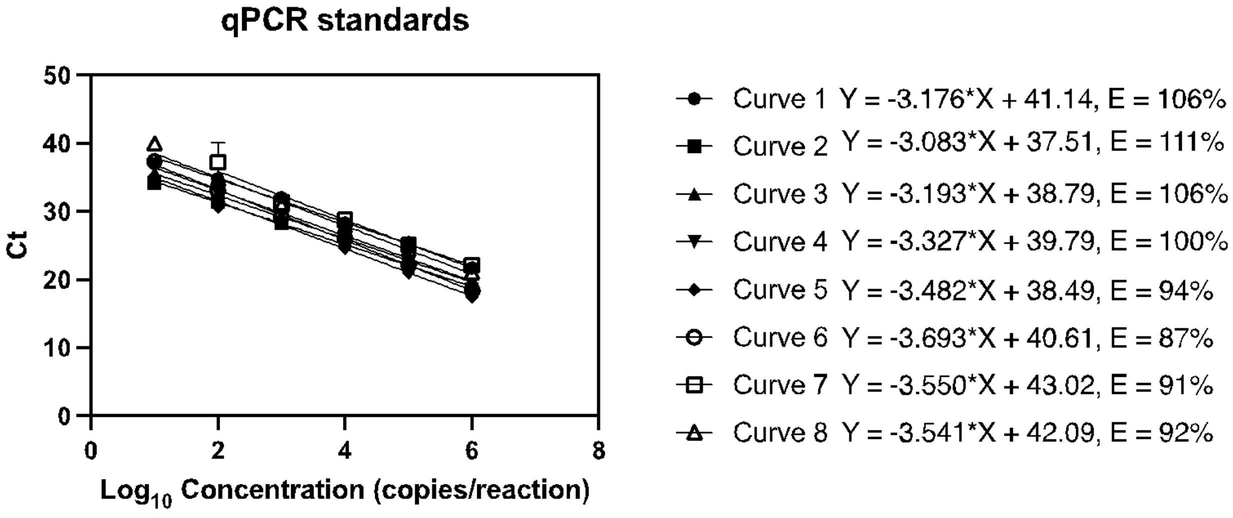


Figure A3: Standard curves for qPCR throughout the project.

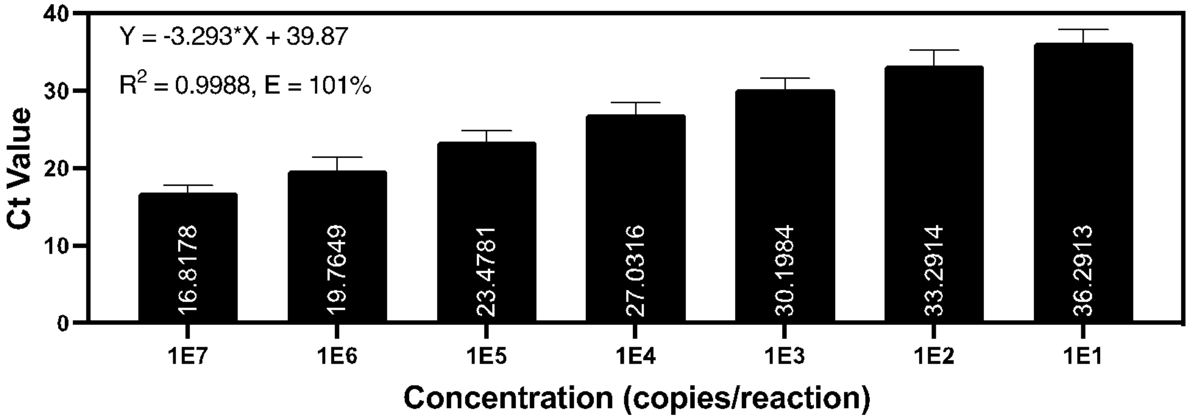


Figure A4: Characteristics of all qPCR standard curves taken together, showing averages for $C_t$ values of each standard concentration.

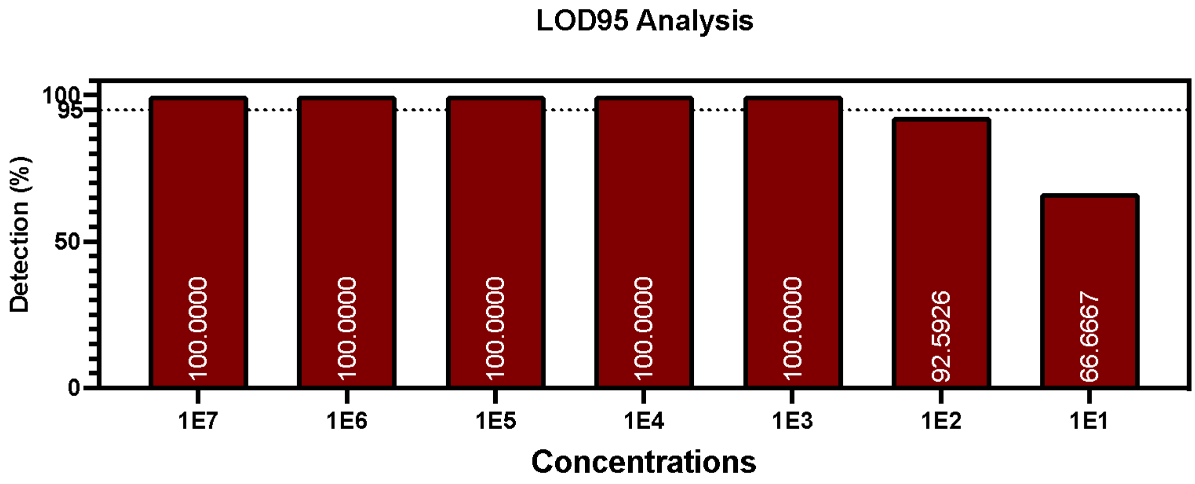


Figure A5: Detection rate (percentages) of all standard curve dilutions throughout the course of the project.

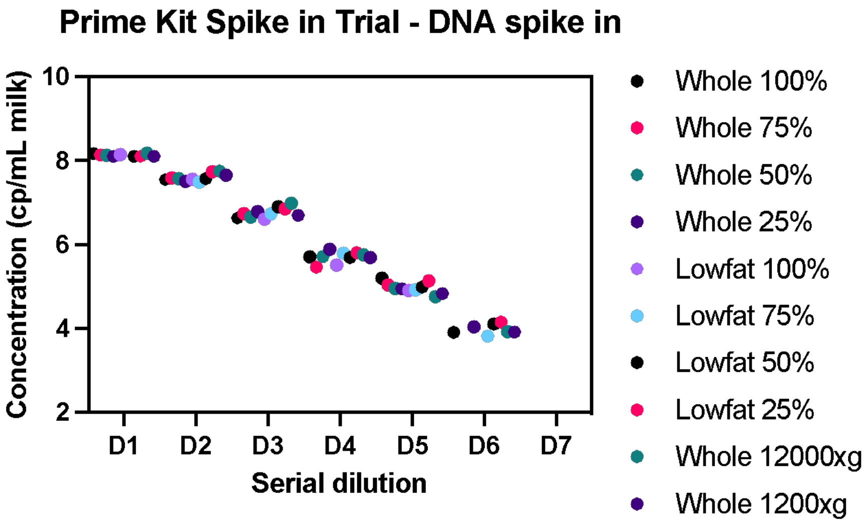


Figure A6: Evaluation of the MagMAX Prime Viral/Total Pathogen NA extraction kit with serial dilutions of spiked in synthetic H5N1 DNA fragments. Milk was diluted with PBS before spike in and two pre-centrifugation conditions were tested (12000xg for 10 minutes and 1200xg for 30 minutes).

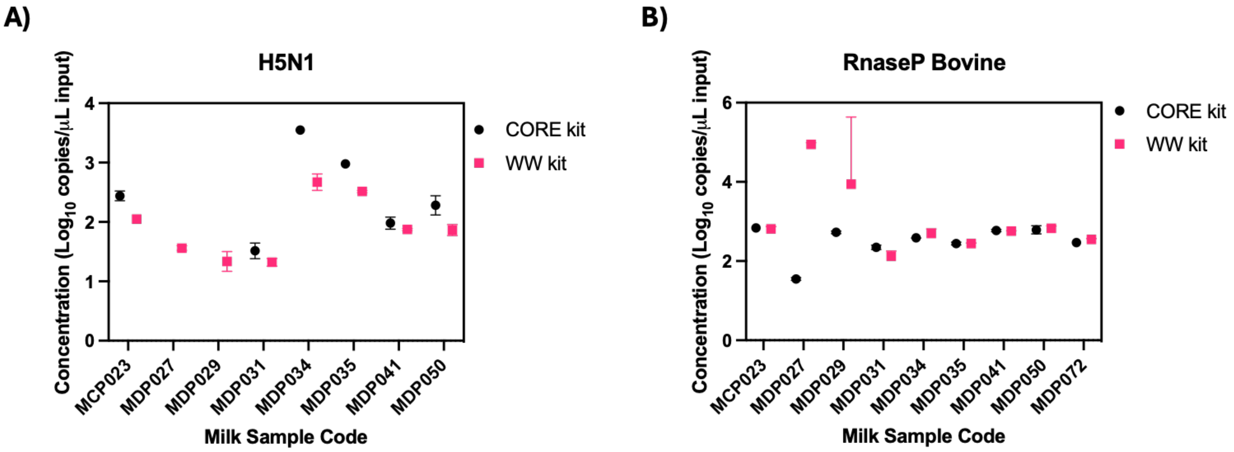


Figure A7: Comparison of the MagMAX CORE extraction kit versus the MagMAX Wastewater extraction kit on a subset of 8 milk samples for A) H5N1 and B) RnaseP Bovine as measured by dPCR.

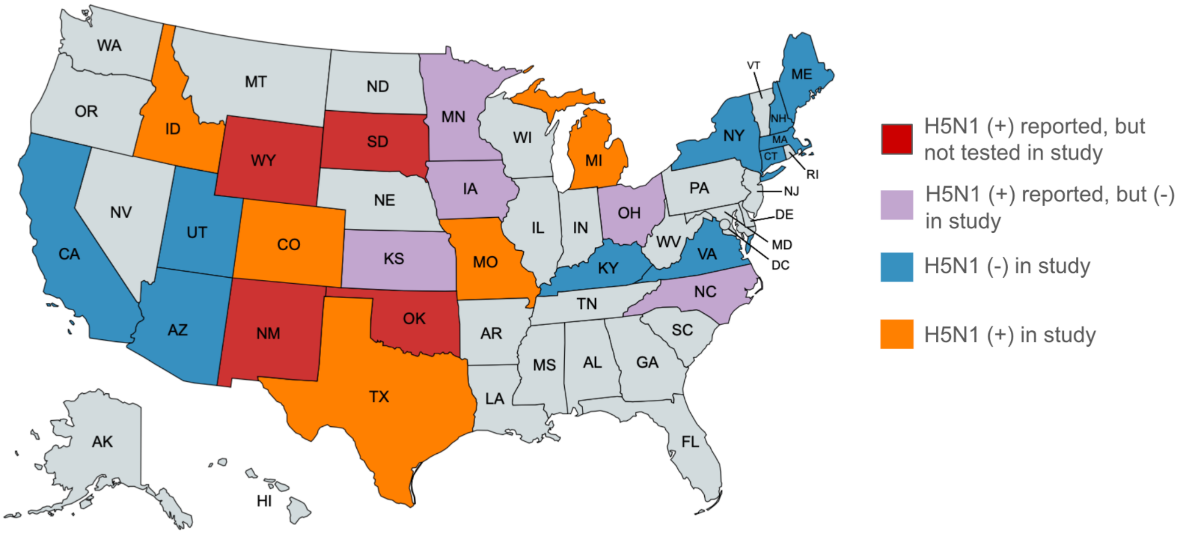


Figure A8: State map depicting states where milk was sourced from for testing in the current study.

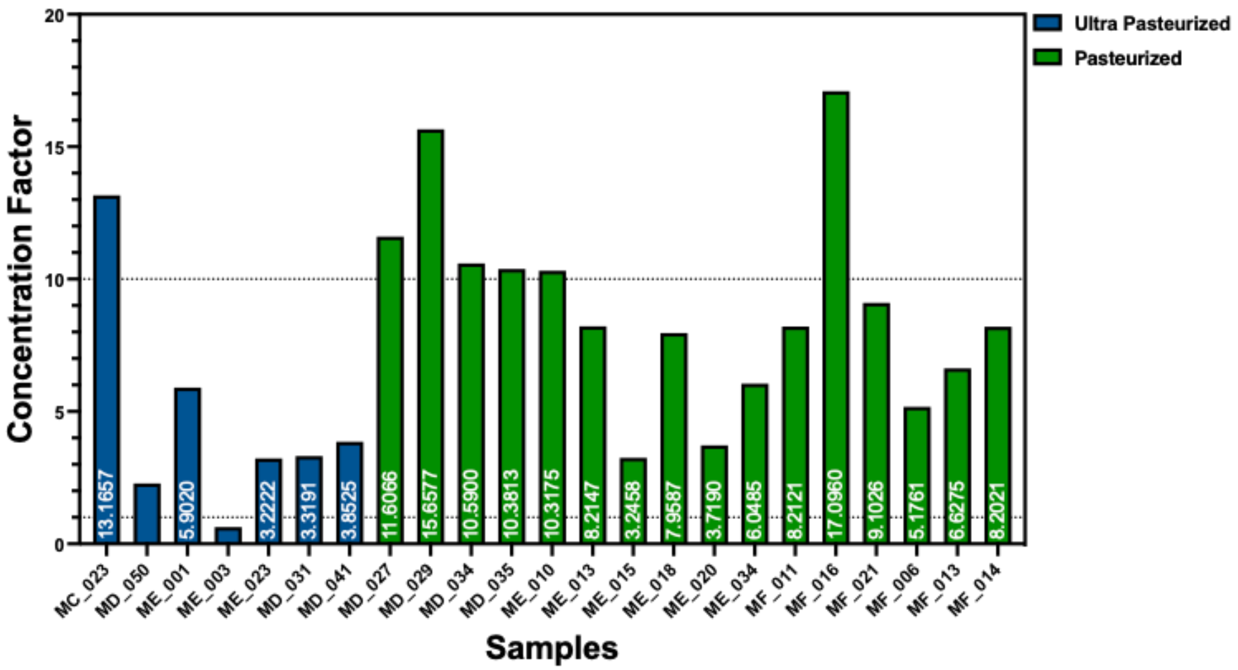


Figure A9: Concentration factor calculated by concentration in a sample after it was extracted ten times and concentrated divided by the initial concentration of the sample.

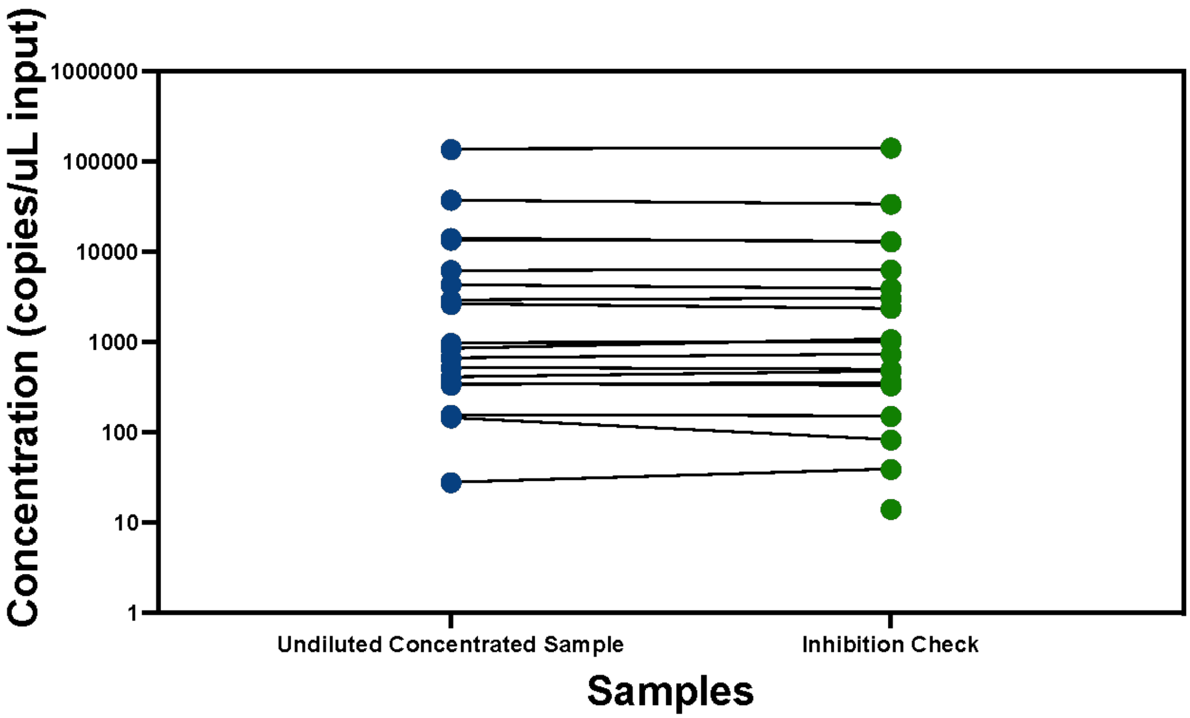

Figure A10: Inhibition check of concentrated samples as measured by dPCR H5_Taq assay. Inhibition check samples received half the amount of template per reaction. Both reactions were normalized to 1µL of input to be able to compare potential effects of PCR inhibitors.

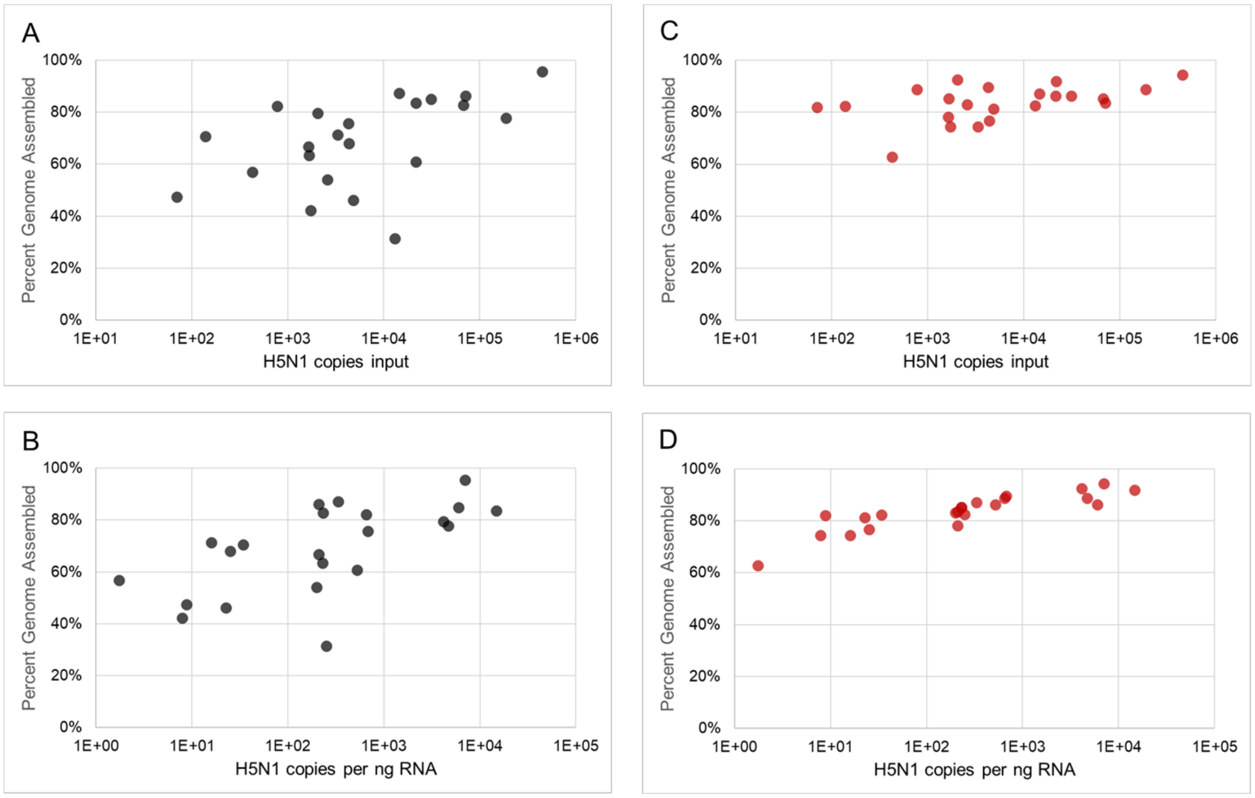


Figure A11: Percent genomes assembled by Amp-Seq using two cycling conditions to generate H5N1 PCR products performed side-by-side on the same cDNA. A) and C) show the data as a function of H5N1 input copies into library construction whereas B) and D) show the same data as a function of H5N1 copies/ng of RNA. Cycling conditions: A) and B): 30s/98°C; 35x (15s/95°C, 5min/65°C); hold at 4°C; (C) and (D): 1min/98°C; 35x (15s/98°C, 30s/65°C, 45s/72°C); 2min/72°C, hold at 4°C.

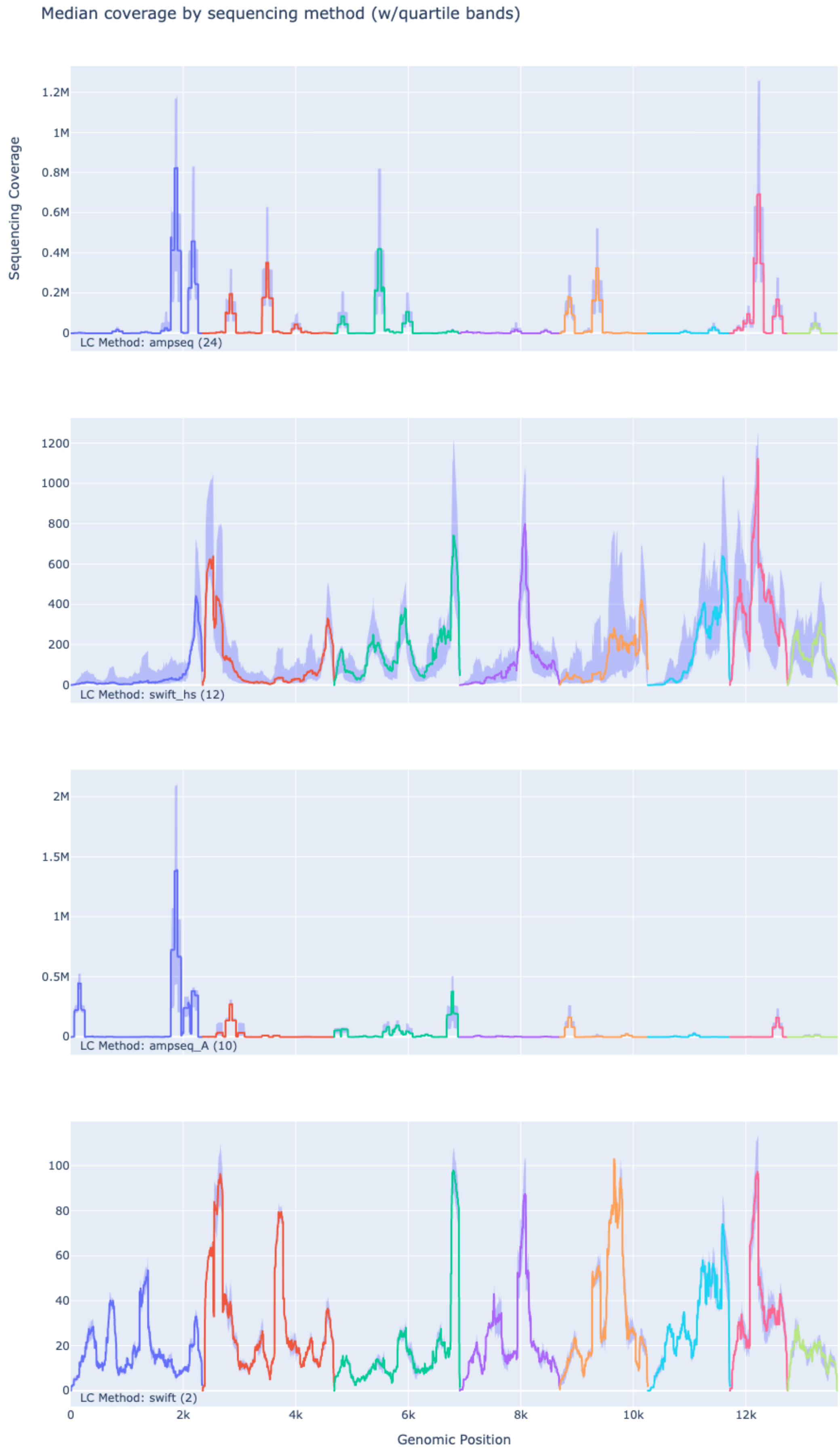

Median coverage by sequencing method (w/quartile bands)
Sequencing Coverage
1.2M
1M
0.8M
0.6M
0.4M
0.2M
0
LC Method: ampseq (24)
1200
1000
800
600
400
200
0
LC Method: swift_hs (12)
2M
1.5M
1M
0.5M
0
LC Method: ampseq_A (10)
100
80
60
40
20
0
LC Method: swift (2)
0
2k
4k
6k
8k
10k
12k
Genomic Position

Figure A12: Sequencing coverage plots, averaged separately for each sequencing method. For each sequencing method, the median sequencing depth across all samples at each genomic position is plotted with a colored line, and 25% - 75% quartile shaded bands are added around the line. Each influenza A genomic segment is plotted in order from segment 1 through 8, each with a different line color. From top to bottom: ampseq refers to the revised Amp-Seq cycling conditions, swift_hs refers to hybrid selected RNA-Seq, ampseq_A refers to the original PCR cycling conditions reported previously, and swift refers to RNA-Seq.

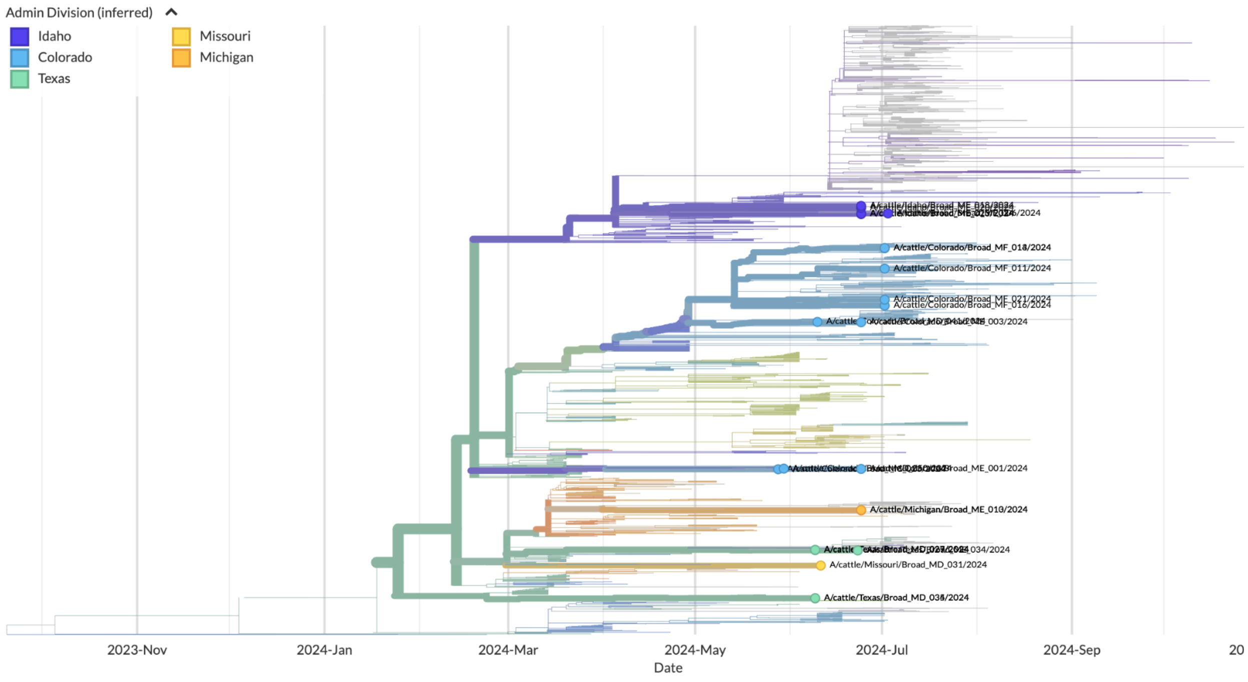


Figure A13: Phylogenetic tree of all cattle-outbreak-associated H5N1-subtype Influenza A concatenated whole genomes from milk samples, as produced by Louise Moncla and the Nextstrain team. Branches and tips are colored by US state of milk processing plant. Tree is shown on a time-axis to reduce the skew caused by some outlier genomes in the contextual data set. See Methods for details. Live build can be found at: https://nextstrain.org/avian-flu/h5n1-cattle-

outbreak/genome?c=division&d=tree,entropy&f_host=Cattle&f_submitting_lab=Broad%20Institute%20Genomic%20Center%20for%20Infectious%20Diseases,%20Genomic%20Center%20for%20Infectious%20Diseases&m=num_date&p=full.

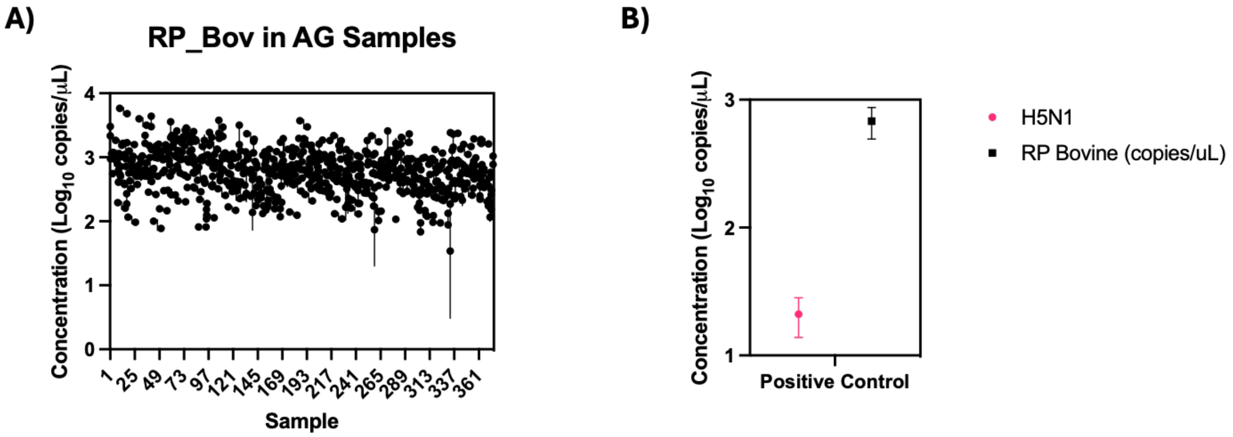


Figure A14: A) Concentration of RnaseP Bovine (RP_Bov) in samples from Massachusetts farms. Note: all samples were negative for presence of H5N1. B) Summary data from positive control milk samples run along with farm samples. Positive control samples were aliquoted from known positive commercial milk samples.

Table A1: PCR primer sequences used in this study.

| Primer | Sequence (5' > 3') | Amplicon length (bp) | Annealing temp | Reference |
|---|---|---|---|---|
| H5_Eva_F | TGCAAACAATTCGACAGAGC | 98 | 58 | This Study |
| H5_Eva_R | GCTTCCCGTTGTGTGTTTTT | | | |
| H5_Taq_F | TATAGARGGAGGATGGCAGG | | | |
| H5_Taq_R | ACDGCCTCAAAYTGAGTGTT | 171 | 59 | Wolfe et al. |
| H5_Taq_P | (FAM)AGGGGAGTGGKTACGCTGCRGAC(IBFQ) | | | |
| RP_Bov_F | CGATTTGGACCTGCGGGCG | 65 | 58 | This Study |
| RP_Bov_R | GAGCAGCGTTCTCCACGAGC | | | |

Table A2: Gene fragment sequences used as positive control, including T7 priming sequence.

| Name | Sequence (5' > 3') |
|---|---|
| H5N1_gblock | gaaatTAATACGACTCACTATAgggACCAGAGGTTGGCACCAAAAATAGCTACTAGATCCCAAGTAAACGGGCA<br>ACGTGGAAGAATGGACTTCTTCTGGACAATCTTAAAACCAGATGATGCAATCCATTTCGAGAGTAATGGAAAT<br>TTCATTGCTCCAGAATATGCATACAAAATTGTCAAGAAAGGGGACTCAACAATTATGAAAAGTGGAGTGGAAT<br>ATGGCCACTGCAACACCAAATGTCAAACCCCAGTAGGTGCGATAAATTCTAGTATGCCATTCCACAACATAC<br>ATCCTCTCACCATTGGGGAATGCCCCAAATACGTGAAGTCAAACAAGTTGGTCCTTGCGACTGGGCTCAGAA<br>ATAGTCCTCTAAGAGAAAAGAGAAGAAAAAGAGGCCTGTTTGGGGCGATAGCAGGGTTTATAGAGGGAGGA<br>TGGCAGGGAATGGTTGATGGTTGGTATGGGTACCATCATAGCAATGAGCAGGGGAGTGGGTACGCTGCGG<br>ACAAAGAATCCACCCAAAAGGCAATAGATGGAGTTACCAATAAGGTCAACTCAATCATTGACAAAATGAACAC<br>TCAATTTGAGGCAGTTGGAAGGGAGTTTAATAACTTAGAAAGGAGGATAGAGAATTTGAACAAGAAAATGGA<br>AGACGGATTCCTAGATGTCTGGACCTATAATGCTGAACTTCTAGTTCTCATGGAAAACGAGAGGACTCTAGA<br>TTTCCATGATTCAAATGTCAAGAACCTTTACGACAAAGTCAGATTACAGCTTAGGGATAATGCAAAGGAGCTG<br>GGTAACGGCTGTTTCGAATTCTATCACAGATGTGATAATGAATGTATGGAAAGTGTGAGAAATGGGACGTAT<br>GACTACCCTCAGTATTCAGAAGAAGCAAGATTAAAAAGAGAAGAAATAAGCGGAGTGAAATTAGAATCAGTA<br>GGAACTTACCAGATACTGTCAATTTATTCAACAGCGGCAAGTTCCCTAGCACTGGCAATCATGATGGCTGGT<br>CTATCTTTATGGATGTGCTCCAATGGGTCGTTACAGTGCAGAATTTGCATTTAGAAAACACCCTTGTTTCTAC<br>TAGTATGCATCTCGT |
| RP_Bov_gblock | gaaatTAATACGACTCACTATAgggGACTTCAGCATGGCGGTGTTTGCCGATTTGGACCTGCGGGCGGGTTCTG<br>ACCTGAAGGCGCTGCGTGGGCTCGTGGAGAACGCTGCTCACCGTGAGTCTCCCGGGCTTCGCGGGCCCCC<br>GTGCCTGCGCTGTCTCGCTGTCCCTAGGCTGTAGAGCCATGCTCTGGAGAGACCCGGCGGGCCTAGTTCT<br>GGTGTCCTGGGGCCTCCGGCGTGTCCTTGGAAACTGATGCCCCTGCGGTGTTGCTCTGACCCGCGGGAAA<br>CTCGAAAGCACTGGGGAGACGTTACCCAGTCCAGCTCCTTCTGTCTTGGGAATTGAGGAAACTGAGGCCCT<br>GACATGGCGGGGGATCTGCATGGGCCTCACAGCTGATGGATAGAAGAAAACAGGGCTCCCAAGTGTACAC<br>CTACACGAGTGCTT |

Table A3: dPCR cycling conditions for H5_Taq

| Step | Temp (°C) | Time | Cycles |
|---|---|---|---|
| RT | 50 | 40m | 1 |
| RT inactivation | 95 | 2m | 1 |
| Denaturation | 95 | 5s | 40 |
| Annealing | 59 | 30s | |
| Imaging | 500ms exposure, 6 gain | | |

Table A4: dPCR cycling conditions for H5_Eva and RP_Bov run as EvaGreen assays.

| Step | Temp (°C) | Time | Cycles |
|---|---|---|---|
| RT | 50 | 40m | 1 |
| RT Inactivation | 95 | 2m | 1 |
| Denaturation | 95 | 15s | 40 |
| Annealing | 58 | 15s | |
| Extension | 72 | 15s | |
| Cooling | 40 | 5m | 1 |
| Imaging | 200ms exposure, 3 gain | | |

Table A5: qPCR cycling conditions for H5_Taq

| Step | Temp (°C) | Time | Cycles |
|---|---|---|---|
| RT | 48 | 15m | 1 |
| RT inactivation | 95 | 10m | 1 |
| Denaturation | 95 | 15s | 40 |
| Annealing | 59 | 1m | |

Table A6: PCR cycling conditions for generating Amp-Seq libraries. Of note, this includes a shorter annealing step (shortened from 5 min) and the addition of an extension step compared to the originally developed protocol.

| Step | Temp (°C) | Time | Cycles |
| --- | --- | --- | --- |
| Activation | 98 | 1m | 1 |
| Denaturation | 98 | 15s | 35 |
| Annealing | 65 | 30s | |
| Extension | 72 | 45s | |
| Final Extension | 72 | 2m | 1 |

Table A7: Input RNA samples for sequencing and best genome assemblies for them. Samples are in the same order as in Figure 5B, i.e. ranked by the length of the most complete genome assembly. H5N1 copies/ul RNA was determined by dPCR. Total RNA concentration and RIN score were determined electrophoretically on a BioAnalyzer chip. We note that the RIN scoring of this assay is for eukaryotic RNA samples, but the length of prominent rRNA bands suggest that much of the RNA is bacterial. Amp-Seq data were generated for all 23 samples. RNA-Seq or hsRNA-Seq data were generated for 18 samples. No RNA-Seq or hsRNA-Seq was performed on the 5 samples indicated by an asterisk.

| Sample | Sample name | State | Pasteurization | H5N1 copies/µL | Total RNA ng/µL | Copies H5N1 per ng RNA | RIN | Longest assembly (kb) | % of Genome | Sequencing method |
|---|---|---|---|---|---|---|---|---|---|---|
| 1 | ME015 | ID | P | 90001 | 12.81 | 7026 | 2 | 13526 | 99.20% | hsRNA-Seq |
| 2 | ME023 | ID | UP | 4345 | 0.29 | 14780 | 2.4 | 13520 | 99.20% | hsRNA-Seq |
| 3 | MF006 | ID | P | 6253 | 1.04 | 6007 | 2.8 | 13476 | 98.90% | hsRNA-Seq |
| 4 | MF014 | CO | P | 14173 | 67.41 | 210 | 5.5 | 13334 | 97.80% | hsRNA-Seq |
| 5 | MF013 | CO | P | 13492 | 58.3 | 231 | 5.8 | 13311 | 97.70% | hsRNA-Seq |
| 6 | MD034 | TX | P | 37574 | 8.02 | 4686 | 5.4 | 13152 | 96.50% | hsRNA-Seq |
| 7 | ME001 | CO | UP | 857 | 1.27 | 677 | 2.5 | 13050 | 95.70% | RNA-Seq |
| 8 | MD050 | CO | UP | 518 | 2.6 | 199 | 2.2 | 12850 | 94.30% | hsRNA-Seq |
| 9 | MD027 | TX | P | 344 | 43.66 | 8 | 3.5 | 12741 | 93.50% | Amp-Seq |
| 10* | ME020 | ID | P | 407 | 0.1 | 4157 | 2.1 | 12619 | 92.60% | Amp-Seq |
| 11* | ME034 | TX | P | 155 | 0.24 | 650 | 2.4 | 12110 | 88.80% | Amp-Seq |
| 12* | MD031 | MO | UP | 145 | 1.02 | 142 | 2.5 | 12065 | 88.50% | Amp-Seq |
| 13 | MF011 | CO | P | 327 | 1.56 | 210 | 6 | 12044 | 88.40% | hsRNA-Seq |
| 14 | MD035 | TX | P | 4329 | 8.27 | 524 | 5.4 | 12009 | 88.10% | hsRNA-Seq |
| 15 | ME013 | MI | P | 2911 | 8.77 | 332 | 7.2 | 11867 | 87.10% | Amp-Seq |
| 16* | MD041 | CO | UP | 333 | 1.45 | 229 | 2.3 | 11614 | 85.20% | Amp-Seq |
| 17 | MC023 | CO | UP | 2623 | 10.48 | 250 | 2 | 11277 | 82.70% | hsRNA-Seq |
| 18 | ME018 | ID | P | 28 | 0.82 | 34 | 2.1 | 11211 | 82.20% | Amp-Seq |
| 19* | ME003 | CO | UP | 14 | 1.6 | 9 | 2.4 | 11181 | 82.00% | Amp-Seq |
| 20 | MD029 | TX | P | 968 | 42.73 | 23 | 3.2 | 11086 | 81.30% | Amp-Seq |
| 21 | MF016 | CO | P | 86 | 49.36 | 2 | 6.3 | 11024 | 80.90% | Amp-Seq |
| 22 | MF021 | CO | P | 870 | 34.62 | 25 | 6.8 | 10456 | 76.70% | Amp-Seq |
| 23 | ME010 | MI | P | 663 | 41.55 | 16 | 2.3 | 10144 | 74.40% | Amp-Seq |

***Detailed methods for PCR assay design and characterization***

Three PCR primer assays were used in the current study (sequences listed in Table A1). First, an H5N1 assay was designed on a subset of HA sequences from the 2.3.4.4.b clade of H5N1 (referred to as H5_Eva). This assay was only used for initial extraction kit evaluation. All digital PCR (dPCR) reactions were run on a Qiacuity One (5plex, Qiagen), using either 24 or 96 well, 8.5k plates and using either OneStep Advanced EvaGreen Kit (H5_Eva and RP_Bov assays) or the OneStep Advanced Probe Kit (H5_Taq), following the manufacturer's protocol. Final reaction concentrations of primer and probe for the H5_Taq assay were 400nM and 200nM, respectively, with 5µL of extraction used as template. Final reaction concentration of primers for both the H5_Eva and RP_Bov assay was 500nM, with 1µL of extraction used for extraction kit trials and 2µL of extraction used as template for milk sample testing (RP_Bov only). Every dPCR run included at least one no template control (NTC) that was used to set the threshold, as well as a positive control. The positive control was made by *in vitro* transcription of an 1800bp synthetic H5N1 DNA sequence of the HA segment to make an RNA standard (sequences provided in Table A2). Transcribed RNA was purified using RNAClean XP beads (Beckman Coulter) following manufacturer recommendations. Purified transcribed RNA was then quantified using the Invitrogen™Qubit™ RNA High Sensitivity (HS) kit following the manufacturer protocol and refined by dPCR. Cycling and imaging protocols for all assays can be found in the Appendix. After thresholding to the NTC, a sample had to have at least three positive partitions to be considered positive (*13*, *31*). Concentrations of samples were then calculated based on dPCR reported concentration, input volume of extraction, and dilution factor.

All quantitative PCR (qPCR) assays were conducted on a QuantStudio 6 Flex (ThermoFisher) using TaqMan RNA-to-$C_t$ 1-Step Kit (ThermoFisher), following manufacturer's recommendations. All samples were run on qPCR in triplicate, 10µL reactions with the final concentration of H5 primers and probe at 500nM and 250nM, respectively (optimization data and cycling conditions can be found in SI). For milk samples, 1µL of extract was used as a template. A sample was considered positive if 2 out of 3 replicates amplified, and if at least one of the extraction replicates was positive. The amplified Ct values were then averaged for subsequent analysis.

***Detailed methods for extraction kit evaluation***

Three commercially available extraction kits were evaluated for their potential to recover nucleic acid from a milk matrix. All kits chosen were bead-based and high-throughput kits compatible with the KingFisher Flex instrument (ThermoFisher) and evaluated for performance by dPCR targeting H5N1 (H5_Eva) and the Ribonuclease P gene of bovines (RP_Bov). First, the MagMAX Prime kit was tested by spiking serial dilutions ($10^2$ - $10^8$ copies/mL) of an 1800bp synthetic DNA fragment of the HA sequence of the H5N1. To evaluate the effect of the milk matrix on recovery, both whole and low-fat milk were tested and diluted with phosphate-buffered saline (PBS) so that the milk matrix was present at 25-100% before being spiked with target. In addition, two pre-centrifugation conditions (either 12000xg for 10 minutes or 1200xg for 30 minutes) were tested with the 100% whole milk condition. Next, the MagMAX CORE kit was evaluated following the manufacturer's "Simple Workflow" which specifies milk as an input type. For this experiment, whole milk was spiked with a dilution series of H5N1 synthetic DNA fragments ($10^2$ - $10^8$ copies/mL) and either processed directly or pre-centrifuged at 12000xg for 10 minutes. Finally, the CORE kit was tested with serial dilutions ($10^2$ - $10^8$ copies/mL) of H5N1 synthetic RNA fragments spiked into both whole and low-fat milk and measured by the H5_Taq assay.

Finally, the MagMAX Wastewater kit was evaluated head-to-head with the MagMAX CORE kit on a subset of retail milk samples previously identified as positive through evaluation with the CORE kit. For this comparison, all samples were re-extracted with the CORE kit as well as processed with the Wastewater kit on the same day. Both kits were used following the manufacturer's instructions, using 200µL of milk as input into extraction.

***Detailed methods for sequencing library generation***

**Unbiased metagenomic libraries (RNA-Seq).** RNA-Seq libraries were generated by the xGen RNA library prep kit (IDT) with 8-base UDI Primers Plate 1 (IDT). Input RNA volumes were adjusted to not exceed 125ng RNA. Time and temperature of the RNA fragmentation step were modified depending on the RIN score following the manufacturer's guidance for low-quality RNA. RNA-Seq libraries were amplified by 5 (>100ng input RNA) or 8 (<100ng input RNA) PCR cycles.

**Hybrid selected RNA-Seq libraries (hsRNA-Seq).** xGen RNA-Seq libraries were pooled in groups of 2-4 libraries (400-900 ng total per pool) from RNA samples with similar (i.e., same order of magnitude) H5N1 copies/µL. Hybrid selection was performed using the Respiratory Virus Research Capture panel as bait (Twist Biosciences) with the Target Enrichment Standard Hybridization v2 kit (Twist Biosciences) following the manufacturer's protocols. Based on initial trials to determine the minimum number of PCR cycles necessary, libraries for hybrid selection were amplified with 9 or 12 PCR cycles to generate >100ng input RNA. Preparative post-hybrid selection PCR reactions (25µL bead slurry in 100µL 1x HiFi HotStart ReadyMix (Roche)) containing 4µM Illumina P7 and 4µM Illumina P5 primers were run for 12-16 cycles for this study.

**H5N1 Amp-Seq libraries (Amp-Seq).** We used 5µL of RNA as input to generate Amp-Seq libraries irrespective of concentration and H5N1 content. cDNA was generated in 20µL reactions using the Superscript IV kit (Thermofisher) following the manufacturer's protocols. Two subsequent PCR reactions (25µL each) were comprised of 1x HiFi HotStart ReadyMix (Roche), 4µL cDNA, and 1.6 µM of previously developed primer pool 1 or 2 specific to H5N1 (15). We used a modified thermoprofile from the original published protocol, namely shortening the annealing step and adding an extension step (Details can be found in Table A6). PCR products were purified with 0.8 volumes of AmPure XP beads (Beckman Coulter) at 0.8 volumes for RNA-Seq and hsRNA-Seq or 1.5 volumes for Amp-Seq, quantitated by Qubit DNA High Sensitivity kit (Thermo Fisher), and characterized on a dsDNA High Sensitivity Bioanalyzer chip (Agilent) using 1ng of PCR product as input. For Amp-Seq, primer pool 1 and 2 PCR products were combined and Illumina sequencing libraries were generated using scaled-down half-reactions of the NEBNext Ultra™ II DNA Library Prep Kit with multiplex oligos (New England Biolabs), adjusting the adapter dilution to the combined input DNA amounts as recommended by the manufacturer: 1:25 for <5ng; 1:12 for 5-25ng; 1:6 for 25-100ng and no dilution for >100ng) and a simple clean-up with 0.9 volumes AmPure beads before the final PCR amplification (6-10 cycles).

Pooled sequencing libraries were sequenced with paired-end 151-base reads on 300-cycle NextSeq 2000 cartridges (Illumina). Separate sequencing runs of metagenomic or HS_metagenomic libraries contained a 10% PhiX spike-in. Four NextSeq sequencing runs were conducted in total: one with 12 metagenomic libraries (0.65nM), one with a hybrid capture of the aforementioned 12 libraries (0.65nM loading concentration), one with the two amplicon sequencing approaches (0.65nM), and one with a superpool of all of the above methodologies at molar ratios of 0.16 (Amp-Seq) : 0.42 (metagenomic) : 0.42 (HS_metagenomic) for sequencing.

***Detailed methods for genomic analysis***

**Basecalling and demultiplexing.** NextSeq sequencing runs (151bp paired end, with 8bp dual barcodes) were basecalled and demultiplexed using Picard using custom specified read structures to accommodate for the xGen library protocol. The first two sequencing runs (RNA-Seq and hsRNA-Seq libraries) were demultiplexed using read structure 151T8B8B20S131T to skip the first 20 bases of the 2nd read that contains artificial sequences added during the "adaptase" step of the protocol. The last two sequencing runs (including Amp-Seq) were demultiplexed using read structure 34S117T8B8B34S117T to remove all PCR primers from the AVRL H5N1 protocol (the longest primer in that design is 34bp, and they appear at the beginning of each read). This produces hard-trimmed reads containing only target sequences and obviates the need for any post-alignment based trimming during consensus sequence generation.

**Genome assembly.** For each sequencing library, consensus influenza genomes were produced using a standard consensus generation pipeline utilized previously for Ebola, Zika and SARS-CoV-2 genomes (*32-35*). The H5N1 Bovine/texas/24-029328-01/2024 reference genome (PP599462.1 through PP599469.1) was used as the reference for all assemblies. For RNA-Seq and hsRNA-Seq libraries, default parameters for *assemble_refbased* were used. For Amp-Seq libraries, the *min_coverage* parameter was increased to 20 (from default 3; as the reads are non-independent) and *skip_mark_dupes* was set to true (from default false; we skip PCR duplicate removal since all reads are PCR duplicates), similar to previously established methods (*36*). As all reads were hard-trimmed of primers during basecalling, we did not perform any post-alignment trimming with BED files. For each sample sequenced by multiple methods, we utilized the hsRNA-Seq genome if it recovered at least 75% of the genome, and, if not, used the Amp-Seq genome.

**Phylogenetic analysis.** Phylogenetic analysis was performed by releasing successful genomes on NCBI Genbank, allowing it to be automatically incorporated into the Moncla Lab / Nextstrain avian-flu builds for the cattle-associated outbreak. Briefly, this utilizes concatenated genomes for the build (all eight segments combined into a pseudo-chromosome) due to the negligible effects of reassortment at the outbreak timescale, excludes certain outlier genomes, and imposes other build-specific parameters found at https://github.com/nextstrain/avian-flu?tab=readme-ov-file#h5n1-cattle-outbreak-2024. The resulting build can be found at https://nextstrain.org/avian-flu/h5n1-cattle-outbreak/genome?c=division&d=tree,entropy&f_host=Cattle&f_submitting_lab=Broad%20Insti

tute%20Genomic%20Center%20for%20Infectious%20Diseases,%20Genomic%20Center%20for%20Infectious%20Diseases&m=num_date&p=full.

**Data availability.** Sequence data is available at NCBI/INSDC under BioProject PRJNA1134696. This includes BioSamples described under the One Health Enterics package, SRA records for each sequencing replicate and method described above, and Genbank & Assembly records for at most one genome per sample, selected by the criteria described above.

***<u>Suggested guidelines for setting up laboratory testing capacity for H5N1 in milk</u>***

In order to stay ahead of the current outbreak, as well as to prepare for worsening conditions such as human-to-human spread, we encourage labs across the nation to set up capacity for H5N1 testing. Based on the study findings, we are able to provide some recommendations to help jumpstart detection at any molecular testing lab by following the provided guidelines:

1. Select PCR detection platform
    a. Important considerations are equipment availability, throughput requirements, and sensitivity requirements.
2. Order materials
    a. Order the H5_Taq primers and probes as well as the RP_Bov primers. If using a different RT-PCR kit than used in the present study, be careful to adjust cycling conditions based on the specific kit characteristics.
    b. Order a nucleic acid extraction kit compatible with your available lab equipment. Several other extraction kits may be compatible than the ones validated in the present study but each should first be evaluated individually before milk sample surveillance.
3. PCR assay set up and validation
    a. Synthetic gene fragments can be ordered from companies such as Twist Biosciences or IDT. Order the gene fragments with the T7 polymerase promoter site on the 5’ end to be able to transcribe into RNA synthetic fragments.

b. In vitro transcribe the gene fragments to create high titer RNA synthetic standards. Clean up material using AmPure XP beads and quantify using Qubit to dilute to the appropriate concentration. A helpful tool to calculate copy numbers is available at: https://nebiocalculator.neb.com/

c. Verify and refine standard material concentration by dPCR, if available.

d. Create a serial dilution of standard material (ideally from 1E1-1E7 copies/uL for qPCR and between 1E1-1E4 for dPCR).

e. Verify PCR performance on the standard material dilutions.

    i. Ensure acceptable performance metrics are met:

        1. Linearity > 90%,
        2. qPCR efficiency between 90-110%
        3. LOD of 10 copies/µL extract or lower

4. Extraction kit set up and validation

    a. Create contrived positive milk samples by spiking negative milk with a serial dilution of synthetic gene fragments (Ideally test a range of concentrations such as 1E2 - 1E8 copies/mL).

    b. Extract contrived samples according to kit specifications.

    c. Evaluate recovered copy number by PCR.

    d. Ensure acceptable performance metrics are met:

        i. Linearity > 90%,
        ii. Process LOD $\lesssim 10^4$ copies/mL of milk
        iii. Consistent RP_Bov detection across H5 dilutions.

5. Sourcing positive milk samples and further validation of lab methods
    a. Identify current outbreak locations at: https://www.aphis.usda.gov/livestock-poultry-disease/avian/avian-influenza/hpai-detections/hpai-confirmed-cases-livestock
    b. Identify state processing plant codes at: www.whereismymilkfrom.com
    c. Purchase milk at local grocery stores originating from states with active outbreaks. As well, source milk from collaborators living in the same geographic regions as active outbreaks to increase your chances of obtaining positive milk samples.
6. Raw milk surveillance
    a. First, verify the lab is capable of receiving and processing raw (unpasteurized) milk and that this does not violate current biosafety protocols.
    b. It is suggested to pasteurize raw milk on site before processing to limit worker exposures while retaining genomic material.
        i. Samples can be pasteurized by heating to an internal temperature of 72°C for at least 15 seconds per the USDA protocol (*37*). It is recommended to validate the temperature and holding time for the specific sampling containers that will be received using retail milk before handling raw milk.
        ii. After heating, cool the milk samples on ice before processing through the same workflow for commercially pasteurized milk.
7. Sequencing and Data Reporting

    a. For low concentration samples (below 500 H5N1 copies/uL extract), optionally re-extract positive samples up to 10 times. Concentrate extracts using a commercially available kit such as the Zymo Clean and Concentrator Kit or another preferred method that includes DNaseI digestion to remove DNA.
    b. For samples with > 500 H5N1 copies/uL, hybrid-selected metagenomic sequencing is suggested to result in more complete genome assemblies. For samples < 500 H5N1 copies/uL, we suggest using an Amp-Seq protocol, such as the one used in the current study.
    c. Deposit raw sequencing reads to GenBank as well as submit assembled genomes to NextStrain.

8. Large Scale Testing Considerations
    a. Depending on scope of the outbreak and number of samples to be processed, consider if a partner lab is needed for scaled up testing.
    b. Partner labs could allow for rapid transition of testing based on needs.
